# Using Family History Data to Improve the Power of Association Studies: Application to Cancer in UK Biobank

**DOI:** 10.1101/2024.07.01.24309759

**Authors:** Naomi Wilcox, Jonathan P. Tyrer, Joe Dennis, Xin Yang, John R. B. Perry, Eugene J. Gardner, Douglas F. Easton

**Affiliations:** Centre for Cancer Genetic Epidemiology, Department of Public Health and Primary Care. University of Cambridge. UK; Metabolic Research Laboratory, Wellcome-MRC Institute of Metabolic Science. University of Cambridge. UK; MRC Epidemiology Unit, Wellcome-MRC Institute of Metabolic Science. University of Cambridge. UK; Centre for Cancer Genetic Epidemiology, Department of Oncology. University of Cambridge. UK

**Keywords:** Power calculations, Family history, Exome Sequencing, Cancer, UK Biobank, Protein-truncating variants

## Abstract

In large cohort studies the number of unaffected individuals outnumbers the number of affected individuals, and the power can be low to detect associations for outcomes with low prevalence. We consider how including recorded family history in regression models increases the power to detect associations between genetic variants and disease risk. We show theoretically and using Monte-Carlo simulations that including family history of the disease, with a weighting of 0.5 compared to true cases, increases the power to detect associations. This is a powerful approach for detecting variants with moderate effects, but for larger effect sizes a weighting of >0.5 can be more powerful. We illustrate this both for common variants and for exome sequencing data for over 400,000 individuals in UK Biobank to evaluate the association between the burden of protein-truncating variants in genes and risk for 4 cancer types.

## Introduction

Genome-wide association studies (GWAS) have been highly successful in identifying common variants associated with disease. Increasingly, association studies are being extended to study rare variants using next-generation sequencing methods. Variants strongly predisposing to disease tend to be maintained at low allele frequencies in the population due to purifying selection, meaning that the power to detect rare variants individually is low, even in very large datasets such as UK Biobank, and careful consideration must be given to analysis methods. A common approach to improve power is to aggregate rare variants, e.g., within genes, using burden tests, with the rationale that similar variants in the same gene are likely to have similar effects. A valuable additional source of information, often collected in research studies, is the family history of a disease. Individuals with affected relatives are more likely to carry risk variants than those without a family history. This phenomenon has been used to design more efficient GWAS by selecting cases enriched for family history (Antoniou & Easton, 2003) and the same argument applies to rare variant association studies. Here we consider how such family history information should be best incorporated in rare variant association analyses. This is particularly relevant in the analysis of data from large cohort studies, in which the number of unaffected individuals far outnumber the cases and the number of unaffected individuals with a family history may be significant. For example, for diseases that are largely restricted to one sex (e.g., breast or prostate cancer), incorporating family history information allows data for individuals of both sexes to be utilised. Utilising ‘proxy’ family history data may also be important in situations where genotypes on cases cannot be obtained (e.g., diseases with high fatality).

Lui et al. (2017) developed a method to test for associations between SNPs and disease by including family history information in controls. They defined ‘proxy cases’ as controls with affected first-degree relatives, and ‘true controls’ to be controls with no affected first-degree relatives. They tested for association between carrier status and the three possible outcomes (case, ‘proxy case’ or ‘true control’) using a 3×2 chi-square test with three outcomes. This test showed greater power to detect associations than a 2×2 chi-square test comparing just cases and controls (Liu et al., 2017). An alternative method, which included information for first-degree relatives of cases as well as controls, was suggested by Hujoel et al. (2019); the Liability Threshold Family History (LT-FH) method. This has greater power than the Liu et al (2017) approach (Hujoel et al., 2019) and has also been used in a rare variant burden test framework: the family history aggregation unit-based test (FHAT). This approach requires the full genotype matrix for each gene to calculate the test statistic (Wang et al., 2022). Our method has a similar rationale, using kinship and first-degree relatives, but uses logistic regression directly on the aggregated burden variable.

In our approach, we consider a logistic regression model to test for an association between disease and gene variant burden that incorporates information on the family history of both cases and controls. Our proposed method weights the disease status of first-degree relatives by k, i.e.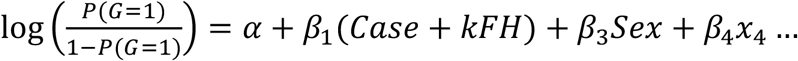 where G is the gene burden, Case is the proband’s disease status (0 or 1) and FH i is the (first-degree) family history of disease status (0 or 1). We consider specifically the case k=1/2, which is theoretically optimal under limiting conditions. We illustrate the method using data from the UK Biobank to show how it improves power to detect associations for several cancers.

## Methods

### Model Motivation

We first consider association tests for a common variant. Let *p* be the frequency of the risk allele, which we assume is associated with an increased relative risk *e*^*β*^ per allele, where *β* is assumed to be small. Let *<u>D</u>*_*i*_ = (*D*_*i*0_, *D*_*i*1_,. *D*_*im*(*i*)_) be the disease phenotypes (0,1) of the j=1..m(i) individuals in family *i=1*..*n*, where individual j=0 is the typed proband, and *<u>G</u>*_*i*_ = (*G*_*i*0_, *G*_*i*1_,. *G*_*im*(*i*)_) be the corresponding genotypes (*G*_*ij*._ = 0, 1 or 2 observed alleles). We show in Supplementary Methods that the score test for *H*_0_: *β*=0 has the form:

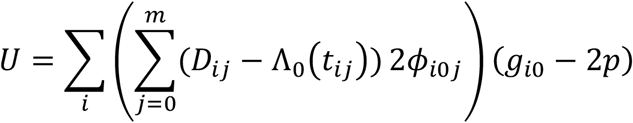

With variance under the null:

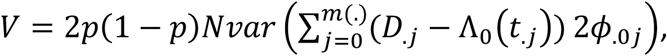

Here *ϕ*_*ijk*_ is the kinship coefficient between individuals *j* and *k* in pedigree *I* and Λ_0_(*t*) is the cumulative disease hazard in the population to age t. Thus, the (locally) most powerful test, for small effect sizes *β*, generalises a simple case-control analysis by replacing the disease status with the disease number of affected individuals in the pedigree, weighted by their degree of relationship to the proband, minus the weighted sum of the predicted number of affected individuals based on population incidence rates, and regressing this against the genotypes.

In practice, full pedigree data are not typically available: it is more usual for summary variables indicating a positive family history (yes/no) or the number of affected relatives, typically just first-degree relatives, to be provided. If we assume that the disease phenotype is relatively rare so that most individuals are unaffected, then the Λ_0_(*t*_*ik*_) terms can be ignored, and the test reduces to regressing the genotypes against 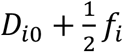, i.e., the disease status of the proband plus ½ the number of affected first-degree relatives *f*. In the analyses considered here, the possibility of more than 1 affected relative is ignored so that *f*_*i*_ = 0 or 1.

### Gene burden tests

Single variant association tests are generally underpowered for rare variants; however, burden tests, in which variants within units, e.g., a gene, are collapsed together can be more powerful if the variants have similar effect sizes (Lee et al., 2014). Here we consider the simplest type of burden test where genotypes are collapsed to a 0/1 variable based on whether samples carry a variant of a given class (e.g., PTVs or rare missense variants in a specific gene). That is, *G*_*i*_ = 1 *if* 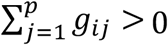 *and* 0 *if* 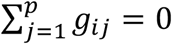 where *g*_*i*._ = 0, 1, 2 is the number of minor alleles observed for sample i at variant j, and p is the number of variants in the gene.

To apply the approach described above, we fit logistic regression models in which the carrier status is the outcome variable, and the disease phenotype is a covariate. Thus, a standard logistic regression model for the case-control study would be of the form:

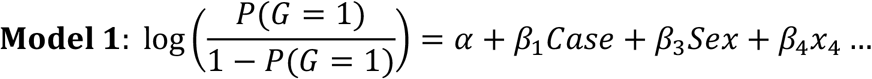

Case is a binary variable for if an individual is a case or control, Sex is a binary variable for female or male, and *x*_3_…represent other covariates, e.g., genetic principal components (PCs). G is a binary variable summarising the genotype (G=1 for a carrier, G=0 for a non-carrier). To incorporate family history, we could add the covariate FH:

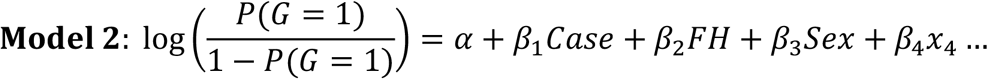

FH is a binary variable for whether the individual has an affected first-degree relative or not. A test for an association is a test of the null hypothesis *H*_0_: *β*_1_ = *β*_1_ = 0, leading to a 2-degree freedom (df) test. The test proposed by Liu *et al* is a *χ*^2^score test, without additional covariates, but a likelihood ratio or Wald test is equivalent.

However, motivated by the arguments above, we consider a modification in which the effect size associated with family history is a predetermined fixed multiple of the case-control effect size, i.e., *β*_2_ = *kβ*_1_.

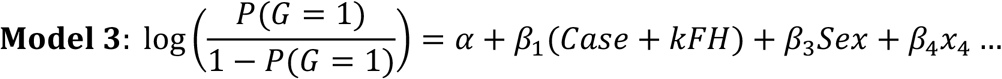

This leads to a 1-df test *H*_0_: *β*_1_ = 0. As above, we postulate that *k*=1/2 is likely to be a reasonable choice. Model 1 is equivalent to k=0, and k=1 is equivalent to treating a positive family history as equivalent to a case.

For the main burden association results, generated using Model 3, we test for association using the Wald p-value associated with *β*_1_. However, to compare the power of Models 1-3 we use the likelihood ratio tests, comparing with the null model:

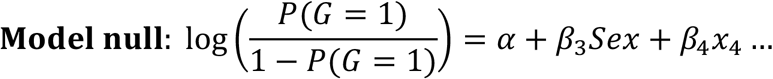

In the case of rare alleles conferring a moderate risk, so that *β* is no longer close to zero, it is less clear that model 3 with k=1/2 provides the most powerful test. As an alternative motivation, we note that for a rare risk allele conferring a relative risk *e*^*β*^, the allele frequency in individuals with an affected first-degree relative is 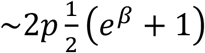 that is approximately the mean of the frequency in cases and controls (Risch, 1990). This corresponds to a model in which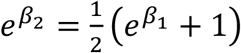. When *β* = *β*_1_ is small this reduces to 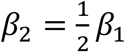 i.e., *k*=½ as proposed. However, for larger effect sizes this predicts 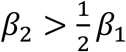; hence, *k*>1/2 may provide a more powerful test in this scenario. We examine the power of alternative values of *k*.

For commoner variants (such as SNPs in GWAS), the genotype *G* has three levels (0, 1 and 2). The equivalent to models 1-3 are then adjacent-categories models:

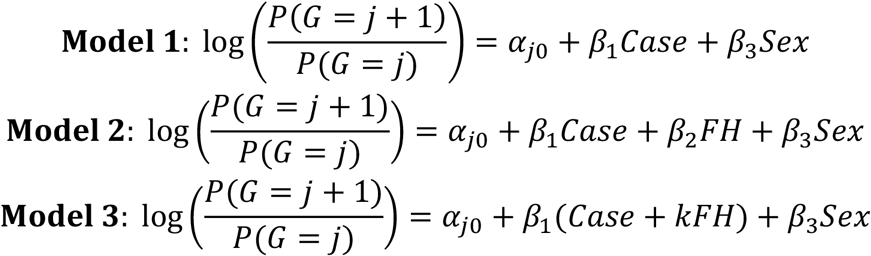

In these models, *β*_1_ is the per-allele OR for the disease associated with the variant, and *β*_2_ is the perallele OR for a positive family history.

### Theoretical Power and Effective Sample Size

We can also derive approximate expressions for the gain in power achievable by incorporating family history in this way, expressed in terms of the relative sample size relative to a case-control study with equal numbers of cases and controls. Suppose there are *N*_0_ controls, *n*_1_ with a family history and *n*_0_ without, and *N*_1_ cases, and n_3_ with a family history and *n*_2_ without.

|  | Control | Case |
| --- | --- | --- |
| **FH=0** | $n_0$ | $n_2$ |
| **FH=1** | $n_1$ | $n_3$ |
| | $N_0$ | $N_1$ |

We let *D*_*ij*_ = 0,1 be the disease status of individual j in family i, as above. Here *d*_*i*_ = *D*_*i*0_ is the disease status of the proband, and *f*_*i*_ = 0,1 according to whether the proband has a positive family history or not. Let *p* be the population allele frequency and *β* the per-allele rate ratio.

In the simple case-control analysis, excluding family history, the standard test is of the form:

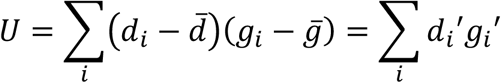

With variance: *V* = *Nvar(<u>d</u>*′)*var*(*<u>g</u>*′)

Where 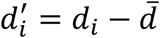 and 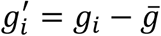 are normalised phenotypes and genotypes with mean 0, and *N* = *N*_0_ + *N*_1_ is the total number of genotyped individuals This gives a Z-score of the form:

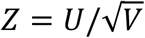

where, under the alternative (see Supplementary Methods):

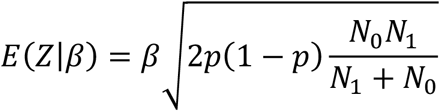

This is a standard formula for deriving the power of a case-control study, excluding family history. The effective sample size is, by definition, the sample size of a case-control study with equal numbers of cases and controls that would give the same power, leading the usual formula:

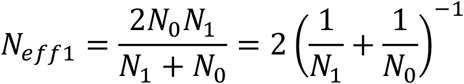

Now we consider the test for our models, which is instead based on:

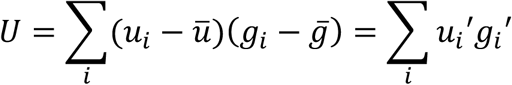

Where the phenotype 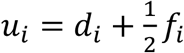, with family history weighted by ½.

We show that (Supplementary Material Methods):

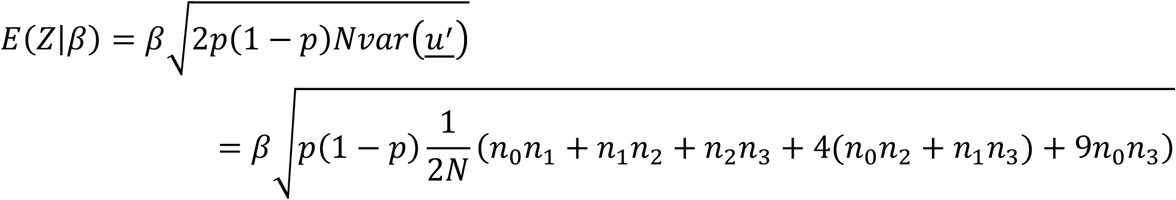

Therefore, the effective sample size is:

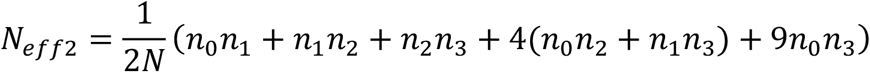

In the absence of family history, i.e. *n*_1_ = *n*_3_ = 0, this reduces to 2*n*_0_*n*_2_/*N* as expected.

It is possible to also calculate an effective population size if instead, the approach of Liu et al (2017) of grouping cases and controls with a family history (“proxy cases”) together, is used. Here the phenotype *y*_*i*_ = max (*d*_*i*_, *f*_*i*_) (Liu et al., 2017).

In this case, the test statistic is of the form:

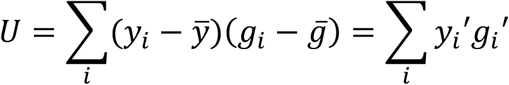

With variance:

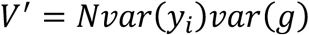

We show that (Supplementary Material Methods):

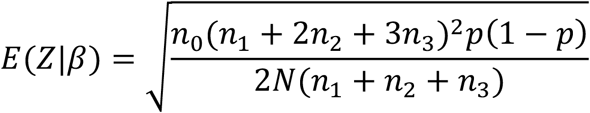

Therefore, the effective sample size is:

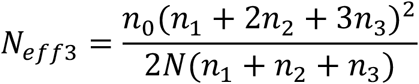

We derive effective sample sizes in UK Biobank for breast, prostate, bowel, and lung cancer using these different approaches.

### UK Biobank

UK Biobank is a population-based prospective cohort study of more than 500,000 individuals. More detailed information on the UK Biobank is given elsewhere (Collins, 2012; Sudlow et al., 2015). WES data for 450,000 samples were released in October 2021 and accessed via the UK Biobank DNA Nexus platform (Backman et al., 2021). QC metrics were applied to Variant Call Format (VCF) files as described by Gardner et Al, including genotype level filters for depth and genotype quality (Gardner et al., 2022). Other filters including samples with disagreement between genetically determined and self-reported sex, excess relatives etc were applied as described elsewhere (Wilcox et al., 2023). The final dataset for analysis included 419,307 samples with 227,393 females and 191,914 males.

Cases for breast cancer, prostate cancer, bowel cancer, and lung cancer were determined by linkage to national cancer registration data (NCRAS) and selecting the appropriate ICD 10 codes (Supplementary Table 1). For breast cancer, we also included self-reported cancer. Both prevalent and incident cases were included. Only cancers which were an individual’s first or second diagnosed cancer were included as cases. The numbers of cases for males and females for each cancer are provided in Table 1.

**Table 1.** The number of female and male cases and controls for each cancer using the ICD-10 codes in Supplementary Table 1.

|  | Females |  | Males |  |
| --- | --- | --- | --- | --- |
|  | Control | Case | Control | Case |
| **Breast cancer** | 209,435 | 17,958 | 191,820 | 94 |
| **Prostate cancer** | 227,393 | 0 | 180,249 | 11,665 |
| **Bowel cancer** | 224,404 | 2,989 | 187,956 | 3,958 |
| **Lung cancer** | 225,641 | 1,752 | 190,009 | 1,905 |

The Ensembl Variant Effect Predictor (VEP) was used to annotate variants, including the 1000 genomes phase 3 allele frequency, sequence ontology variant consequences and exon/intron number (McLaren et al., 2016). Annotation files were used to identify PTVs and rare (allele frequency <0.001 in both the 1000 genomes dataset and the current dataset) missense variants. PTVs in the last exon of each gene and the last 50 bp of the penultimate exon were excluded as these are generally predicted to escape Nonsense-Mediated mRNA Decay (NMD).

UK Biobank imputed genetic data was also accessed to explore results for 4 known breast cancer GWAS SNPs (Easton et al., 2007; Fachal et al., 2020; Michailidou et al., 2017; Turnbull et al., 2010). Genome-wide genotyping was performed using the UK Biobank Axiom Array, where approximately 850,000 variants were measured, and this was imputed to >90 million variants using the Haplotype Reference Consortium and UK10K+10000 Genome’s reference panels. This dataset contained information for 451,959 individuals (245,215 females).

### Monte Carlo Simulations

We used Monte Carlo simulations to compare the power of models 2 and 3. We simulated N datasets under alternate hypotheses with specified OR and k values, each of size n=450,000. The generated proportions of males and females, cases/controls and positive and negative family history were the same as for cancer phenotypes in the UK Biobank dataset (Table 2). For each variable, this was done using random sampling from a binomial distribution. We then generated a binary carrier variable, indicating if an individual carried a risk variant, based on predicted probabilities from the logistic regression with coefficients (*α, β*_1_=log(OR), *β*_2_ = klog(O*R*), *β*_3_). This was repeated for each of the 5,000 simulated datasets. The power was calculated as the proportion of times the null hypothesis was correctly rejected for the models, i.e., the proportion of p-values less than 0.05 for nominal significance, or less than 2.5×10^−6^ for exome-wide significance, based on the appropriate likelihood ratio test.

**Table 2.** Proportions used to simulate datasets for MC power calculations for the 4 cancers.

|  | Females | Cases within Females | Cases within Males | FH=1 within Female Controls | FH Cases within Female Cases | FH Cases within Male Controls | FH Cases within Male Cases |
| --- | --- | --- | --- | --- | --- | --- | --- |
| **Breast Cancer** | 0.54 | 0.079 | 0.00049 | 0.11 | 0.18 | 0.10 | 0.16 |
| **Prostate Cancer** | 0.54 | 0 | 0.061 | 0.081 | 0 | 0.077 | 0.15 |
| **Bowel Cancer** | 0.54 | 0.013 | 0.021 | 0.11 | 0.15 | 0.12 | 0.18 |
| **Lung cancer** | 0.54 | 0.0077 | 0.0099 | 0.13 | 0.23 | 0.13 | 0.22 |

## Results

### Breast Cancer: known genes

The ORs and p-values for the five “known” genes, under the different models, are given in Table 3. The association was most significant for model 3 (assuming k=1/2) for truncating variants in *ATM, CHEK2* and *PALB2*, and also for rare missense variants in *CHEK2*. For *BRCA1* and *BRCA2*, the association tests were more significant under model 2. It is notable that for these genes, which are associated with the highest risks, the estimated values of k were the largest (all greater than 0.5), as expected. However, only for *BRCA1* and *BRCA2* did estimating k improve the significance, showing the benefit of model 3 for detecting associations with modest effect sizes.

**Table 3.**
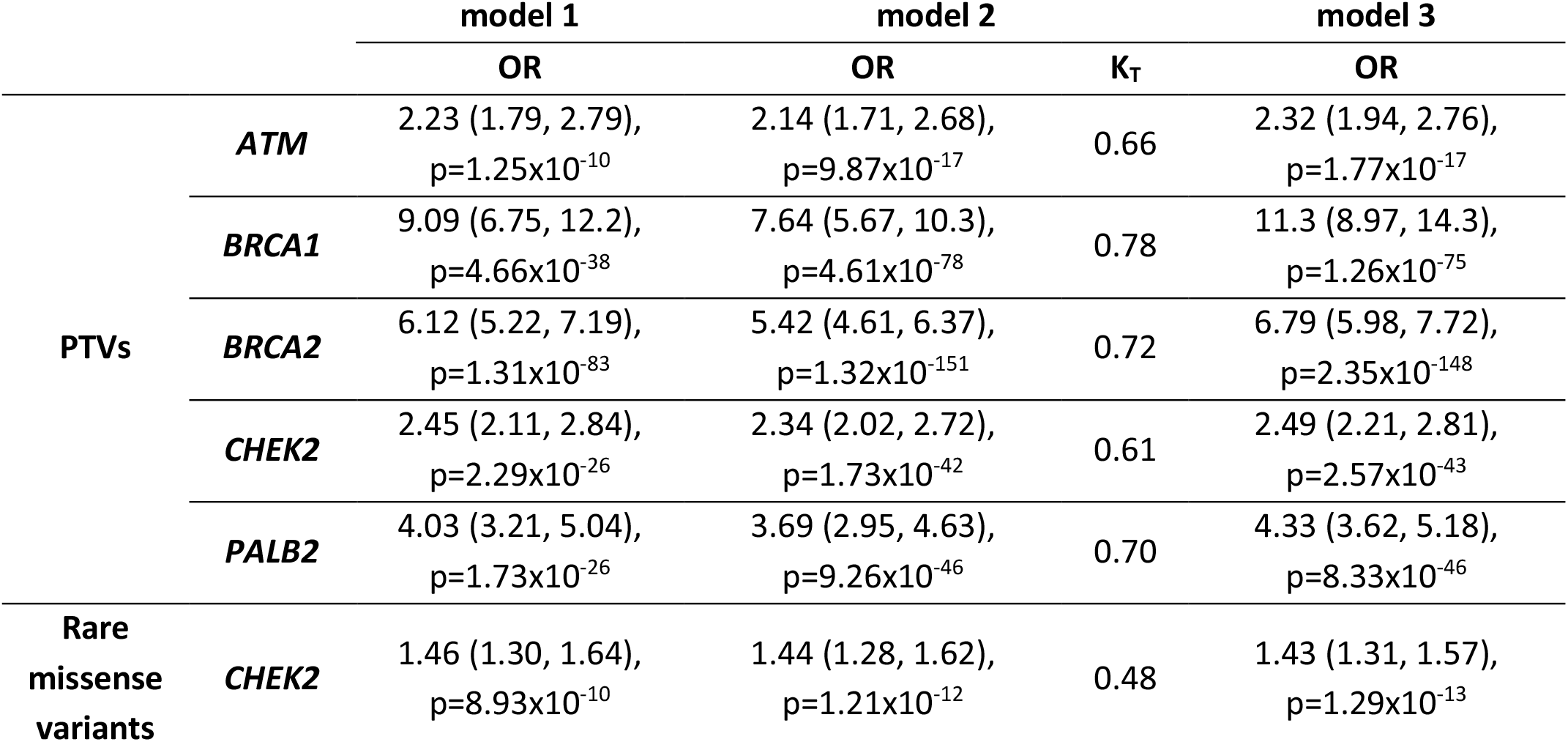
ORs from models 1-3 for breast cancer risk for PTVs in known risk genes. For model 2 the ‘true’ *k*_*T*_ = *β*_2_/*β*_1_ is calculated, and model 3 uses k=1/2. P-values are from the LRT to the Null model.

### Breast cancer GWAS SNPs

Table 4 shows the corresponding results for GWAS SNPs. Here the analyses using model 3 (fixing k=1/2) were consistently the most significant. Under model 2, the best estimates of k are between 0.4 and 0.6, consistent with the theoretical expectation.

**Table 4.**
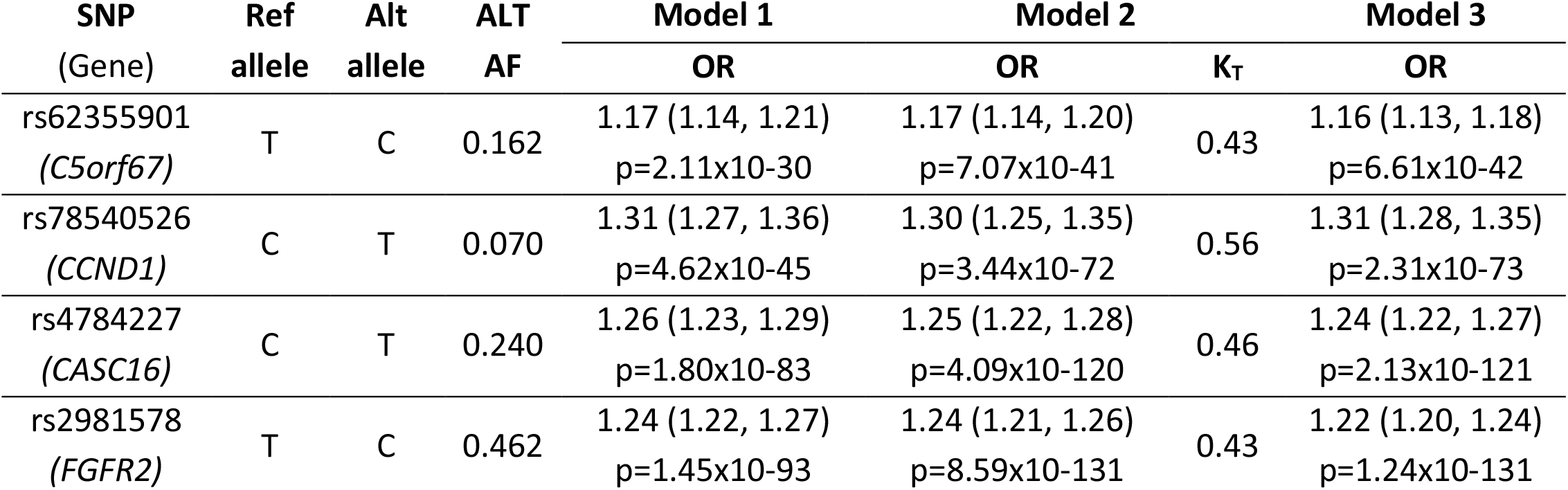
Per-allele ORs from models 1-3 for 4 lead breast cancer GWAS SNPs. For model 2 the ‘true’ *k*_*T*_ = *β*_2_/*β*_1_ is calculated, and model 3 uses k=1/2. P-values are from LRT to the null model.

### Effective sample size

Figure 1 summarises the effective size sample sizes for the analysis of the four cancer types in UK Biobank, using the different methods. As expected, the effective sample size is greatest for breast cancer, reflecting the higher prevalence of this cancer. The relative gain in effective sample size is, however, greatest for lung cancer (more than 3-fold) reflecting that the proportion of individuals with a positive family history is highest for this cancer.

**Figure 1.**
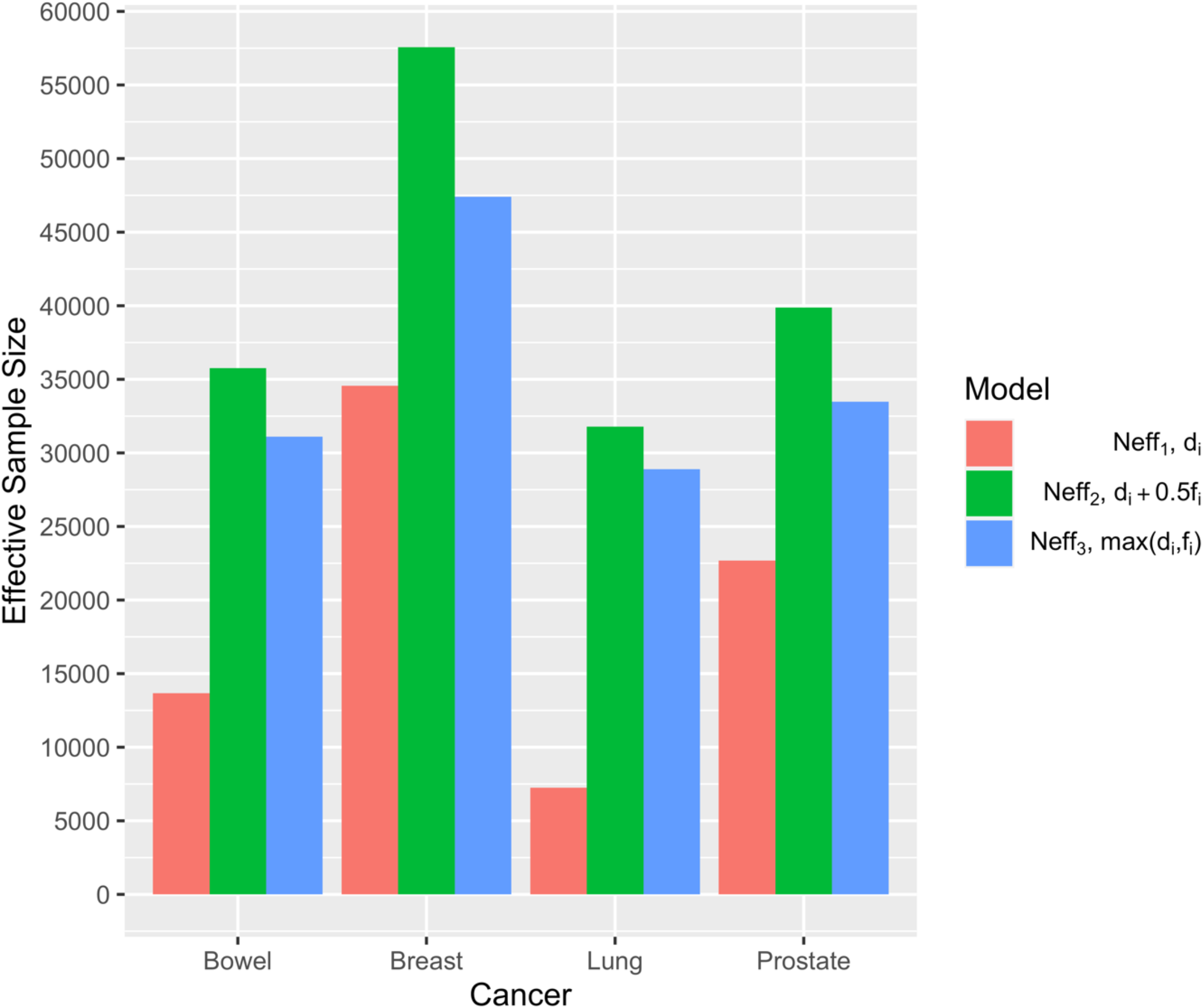
Effective sample size for different models. Neff_1_ is the effective sample size from the standard case-control analysis (Model 1) just considering an individual’s disease status, d_i_. Neff_2_ is the effective sample size from our model including family history, f_i_, of cases and controls with a weighting 0.5 (Model 3). Neff_3_ is the effective sample size from the Model of Liu et al (2017) where controls with a positive family history are treated the same as true cases (Liu et al., 2017).

### Monte Carlo Simulations

We compared the power of model 2 and model 3 by Monte Carlo simulation for the four cancers.

For breast cancer, at a significance level of 0.05, the power is greater for model 3 than model 2 or model 1 for OR<2.5 (Figure 2). For example, the power to detect OR=2 increases from 0.964 to 0.989 to 0.995 when comparing models 1 to 3. For OR≥2.5 the power approaches 1 for all models. At exome-wide significance, the power is also greatest for model 3 for OR<3. For example, the power to detect OR=2 increases from 0.232 to 0.433 to 0.526 comparing models 1 to 3. For OR≥3 the power approaches 1 for all models. Varying the value of *k* between 0.4 and 0.7 under model 3 made little difference to the power for any OR (Supplementary Tables 3 and 4, Supplementary Figure 1).

**Figure 2.**
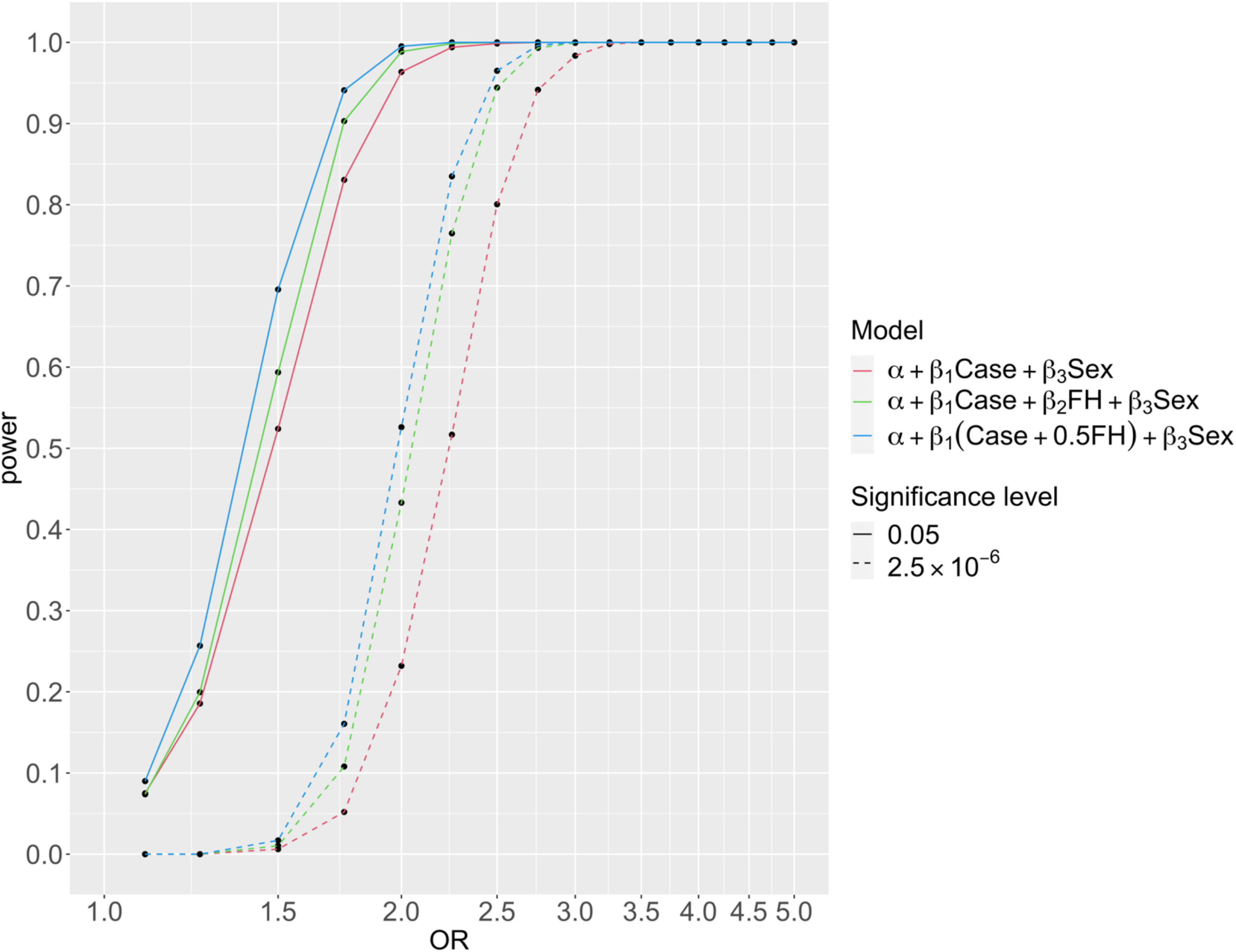
The power to detect different ORs for breast cancer risk. Power is calculated by Monte Carlo simulations for 5,000 datasets of size 450,000 with proportions of sex, case/control, and family history the same as for breast cancer in the UK Biobank. For each simulated dataset model 2 and model 3 were compared to the null model by LRT.

For prostate cancer, the power of each model is lower than that for breast cancer (Supplementary Figs. 2 and 3) reflecting the lower prevalence of the disease. At a significance level of 0.05, the power is greater for model 3 than model 2 or model 1 for OR<2.5 (Supplementary Figure 4). For example, the power to detect an association with OR=2 increases from 0.86 to 0.94 to 0.97 comparing models 1 to 3. For OR≥2.5 the power approaches 1 for all models. At exome-wide significance, the power is also greatest for model 3 for OR<3.5. For lung cancer, the power of each model is lower than that for breast, prostate, and bowel cancer (Supplementary Figs. 2 and 3) reflecting the lower prevalence of the disease. At a significance level of 0.05, the power is greater for model 3 than model 2 or model 1 for OR<3 (Supplementary Figure 5). For example, the power to detect OR=2 increases from 0.48 to 0.87 to 0.92 comparing models 1 to 3. For OR≥3.5 the power approaches 1 for all models. At exome-wide significance, the power is also greatest for model 3 for OR<4. Finally, for bowel cancer, the power of each model is lower than that for breast cancer and prostate cancer, but greater than that for lung cancer (Supplementary Figs. 2 and 3). At a significance level of 0.05, the power is greater for model 3 than model 2 or model 1 for OR< 3 (Supplementary Figure 6).

### Exome-wide association results using Model 3

#### Breast cancer

Exome-wide association analyses for breast cancer using this approach have been presented elsewhere (Wilcox et al., 2023). In brief, for PTVs, 30 genes were associated at P<0.001. Of these, 6 genes reached exome-wide significance (*P*<2.5×10^−6^): the five previously known genes above plus *MAP3K1*. The results in Wilcox *et al*. were based on a meta-analysis of UK Biobank and studies in the Breast Cancer Association Consortium, but the same 6 genes reached exome-wide significance using UK Biobank alone. If the analyses had been based on model 1 in the UK Biobank alone, 23 genes would have met P<0.001 compared to 27 using model 3 (Supplementary Table 5). In UK Biobank, all the p-values for the exome-wide significant genes from model 3 were at least 10^−3^ smaller than using model 1, e.g. for *MAP3K1* the p-value for model 3 was 3.2×10^−8^ compared to 8.3×10^−5^ in model 1 (Supplementary Table 5).

We applied the same approach to three other common cancers for which family history data are available in UK Biobank: prostate, bowel, and lung.

#### Prostate cancer

For prostate cancer, 34 genes were associated at P<0.001 (Supplementary Table 6, Supplementary Figs. 7 and 8). Of these, 3 met exome-wide significance; *BRCA2, CHEK2* and *ATM*. Associations at P<1×10-4 were also identified for PTVs in *GEMIN2, OSGIN1, UBQLN4, C9orf50* and *C9orf152*. There was some evidence of association for *BAP1* (p=0.0010) and *MSH4* (p=0.0018). If the analyses had been based on model 1, 26 genes would have met P<0.001 compared to 34 using model 3. The p-values for the exome-wide significant genes from model 3 were all more significant than model 1, e.g. for *GEMIN2* the p-value for model 3 was 3.9×10^−6^ compared to 3.1×10^−4^ in model 1 (Supplementary Table 6).

#### Lung cancer

For lung cancer, 46 genes were associated at p<0.001 (Supplementary Table 7, Supplementary Figs. 9. and 10). Of these, none met exome-wide significance, but associations at P<1×10^−4^ were observed for *MON2, ASB6, ABCF2, PPP6R3, ARHGAP35, KCNH8* and *BIRC3*. However, we note that the number of case carriers for these genes was very low (≤ 4) and standard errors were large. Of known cancer susceptibility genes, some evidence of association was seen for *ATM* (p=0.00012) and *BRCA2* (p=0.0025). If the analyses had been based on model 1, 56 genes would have met P<0.001 compared to 46 using model 3. However, the p-values for the most significant genes from model 3 were all more significant than model 1, e.g., for *ATM* the p-value in model 3 was 1.2×10^−4^ compared to 2.0×10^−3^ in model 1 (Supplementary Table 7).

#### Bowel cancer

For bowel cancer, 42 genes were associated at P<0.001 (Supplementary Table 8, Supplementary Figs. 11 and 12). Of these, 5 met exome-wide significance: the known susceptibility genes *MSH2, MSH6, MLH1* and *APC;* and *GAPDH*. The mismatch repair gene *PMS2* also showed evidence of association (P=0.00020). Associations at P<1×10-4 were also observed for *MT1G, FLCN, SMAD4* and *ATF3*. Among other cancer susceptibility genes, associations were seen for *ATM* (p=0.0013), *BRCA1* (p=0.0015), *BARD1* (p=0.00021), *CHEK2* (p=0.039), *RAD51D* (p=0.0065), *MSH3* (p=0.0025). If the analyses had been based on model 1, 64 genes would have met P<0.001 compared to 42 using model 3. However, the p-values for the exome-wide significant genes were all more significant than model 1, with no additional genome-wide significant genes using model 1, e.g. for *GAPDH* the p-value in model 3 was 9.3×10^−7^ compared to 1.9×10^−5^ in model 1 (supplementary Table 8).

## Conclusions

Our analyses confirm that the inclusion of family history can improve the power of association studies, for both common variants (the focus of GWAS) and rare variants (the focus of WES studies). We show that for typical common variants, and for “moderate” risk gene variants such as PTVs in *CHEK2* and *ATM*, a 1-degree freedom test fixing (assigning a weight k=1/2 to individuals with a family history) is more powerful than a 2-degree freedom test in which the effects of genotype on disease risk and family history are both estimated. This is consistent with the theoretical results that the 1-degree freedom testing with k=1/2 should be most powerful in the limiting case of small effect sizes.

For variants with larger effect sizes, the association test the 2df can be more powerful (or, equivalently, the 1df test can be made more powerful by assuming a larger k). This is logical: in the simplifying situation with a single relative, the relative risk associated with a positive family history 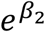, should be approximately 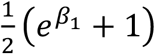. When *β*_1_ is large, 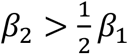, as is observed for *BRCA1, BRCA2* and *PALB2* for breast cancer. In practice, however, we expect that most novel variants will be associated with ORs<2.5, corresponding to 0.5<k<0.6. In theory, the power would be improved by fixing k to its optimum value, but in practice, we do not know k, and k=1/2 is a straightforward choice giving near optimum power. We also note that, in theory, it would be possible to fit the constrained model in which 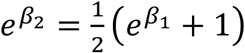 rather than the simpler 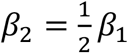 constraint. However, this is a non-linear model which is less suitable for genome-wide analyses involving many genes or variants, and the model would still be approximate (for example, dealing only approximately with cases with a family history).

It should be noted that fixing k=0.5 will tend to overestimate β_1_ if the effect size is large, as seen in the analysis the higher risk known genes. Thus, this is not an ideal approach for estimating risk. However, the focus here is on genome-wide discovery experiments, where the main interest is in the power to detect associations rather than effect size estimation.

While the gain in power was seen for all cancers, it was particularly marked for lung cancer, reflecting the relatively large proportion of affected relatives (in turn related to the much higher risk in older individuals in early birth cohorts).

The gain in power by incorporating family history is illustrated by the exome-wide association analyses for specific cancers. Using model 3 with k=0.5, novel association genes at exome-wide significance were identified for breast cancer e.g., *MAP3K1*, as reported elsewhere (Wilcox et al., 2023). For bowel cancer, we note *MSH2, MSH6, MLH1*, and *PMS2* are MMR genes with known bowel cancer associations, while *APC* is a known susceptibility gene through its association with familial adenomatous polyposis. Biallelic *MSH3* variants have also been associated with adenomatous polyposis. To our knowledge the association for *GAPDH* is novel. Literature suggests *GAPDH* expression to be significantly up-regulated in human colorectal carcinoma tissues compared to adjacent normal tissue (Tang et al., 2012). Genes associated with prostate cancer at exome-wide significance include *BRCA2*, a known risk factor, as well as other breast-cancer risk genes *CHEK2* and *ATM* for which previous evidence has been more equivocal. No significant associations were observed for *BRCA1* or *HOXB13*, previously identified risk genes (Ewing et al., 2012; Nyberg et al., 2020; Nyberg et al., 2019). However, the *BRCA1* association remains controversial, and there were only 12 case carriers in this dataset, while the reported *HOXB13* association is specific to the p.Gly84Glu missense variant not included in these analyses. For lung cancer, no genes reached exome-wide significance (using any model). The genes with more moderate evidence can, however, provide a basis for further targeted replication studies. There were more genes at P<10^−4^ using model 1 than model 3 for lung and bowel cancer (in contrast to the theoretical expectations), however many of these genes had very low carrier counts and this may reflect a combination of chance and inaccuracy in the type I error at very low frequencies.

While we have concentrated on the application of these methods to UK Biobank, the same approach could be fruitfully applied to other cohorts: for example, AllOfUs has family history for a wide range of cancers, as well as other diseases (“The “All of Us” Research Program,” 2019; Bick et al., 2024). Possible further developments to the model, in datasets where the information is available, would include incorporating more extensive family history information e.g., extending the model to 2nd-degree relatives, as well as including data on genotypes of relatives, which would further improve power.

In conclusion, these results demonstrate that including family history in burden regression models improves the power to identify cancer susceptibility genes. This is particularly relevant in the analysis of data from large cohort studies where the number of unaffected individuals outnumbers the number of affected individuals, and the number of unaffected individuals with a family history is significant. We demonstrated the power of this model for 4 cancer phenotypes, but the method could also be applied to non-cancer phenotypes where family history information is available, such as Alzheimer’s disease, major depressive disorder, and coronary artery disease in the UK Biobank.

## Data Availability

All data produced in the present study are available upon reasonable request to the authors

## Acknowledgements

QC of the data has been funded by the Medical Research Council (unit programs: MC_UU_12015/2, MC_UU_00006/2). The research has been conducted using the UKB Resource under application number 28126. N.W. was supported by the International Alliance for Cancer Early Detection, an alliance between Cancer Research UK (C14478/A29329), Canary Center at Stanford University, the University of Cambridge, OHSU Knight Cancer Institute, University College London and the University of Manchester. J.D. was supported by core funding from the NIHR Cambridge Biomedical Research Centre (NIHR203312). X.W. and J.P.T. were supported by Cancer Research UK (PPRPGM-Nov20\100002, PRCPJT-May21\100006 and G110748).

## Author contribution statement

D.F.E. supervised this work and directed the overall analysis. N.W. performed the statistical analysis. N.W., E.J.G., J.P.T., J.D.P. developed the bioinformatics and computational pipelines. X.Y. and J.D. acquired data and X.Y. extracted cancer phenotypes. N.W. and D.F.E. drafted the manuscript. All authors reviewed and approved the paper.

