## Supplementary Material for "Using Family History Data to Improve the Power of Association Studies: Application to Cancer in UK Biobank"

### Supporting Information

### Supplementary Methods

#### Model Motivation

Let $p$ be the frequency of the risk allele, which we assume is associated with an increased relative risk $e^{\beta}$ per allele, where $\beta$ is assumed to be small. Let $\underline{D_i} = (D_{i0}, D_{i1}, .. D_{im(i)})$ be the disease phenotypes (0,1) of the j=1..m(i) individuals in family *i=1..n*, where individual j=0 is the typed proband, and $\underline{G_i} = (G_{i0}, G_{i1}, .. G_{im(i)})$ be the corresponding genotypes ($G_{ij}$ = 0, 1 or 2 observed alleles).

The likelihood for the genotype data of probands, conditional on observed family phenotypes, is of the form:

$$L = \prod_i \frac{P(G_{i0} = g_{i0})P(\underline{D_i}|G_{i0} = g_{i0})}{p^2 P(\underline{D_i}|G_{i0} = 2) + 2p(1-p)P(\underline{D_i}|G_{i0} = 1) + (1-p)^2 P(\underline{D_i}|G_{i0} = 0)}$$

Therefore:

$$\log(L) = \sum_i \log\left(\frac{P(G_{i0} = g_{i0})P(\underline{D_i}|G_{i0} = g_{i0})}{p^2 P(\underline{D_i}|G_{i0} = 2) + 2p(1-p)P(\underline{D_i}|G_{i0} = 1) + (1-p)^2 P(\underline{D_i}|G_{i0} = 0)}\right) = \sum_i \log(f(\beta))$$

Where $f(\beta) = \frac{P(G_{i0} = g_{i0})P(\underline{D_i}|G_{i0} = g_{i0})}{p^2 P(\underline{D_i}|G_{i0} = 2) + 2p(1-p)P(\underline{D_i}|G_{i0} = 1) + (1-p)^2 P(\underline{D_i}|G_{i0} = 0)}$

The first derivative of the likelihood is:

$$\frac{dlogL}{d\beta} = \sum_i \frac{d}{d\beta}\log(f(\beta)) = \sum_i \frac{1}{f(\beta)}\frac{df(\beta)}{d\beta}$$

$$= \sum_i \frac{dP(\underline{D_i}|G_{i0} = g_{i0})}{P(\underline{D_i}|G_{i0} = g_{i0})d\beta} - \frac{\left[p^2 \frac{dP(\underline{D_i}|G_{i0} = 2)}{d\beta} + 2p(1-p)\frac{dP(\underline{D_i}|G_{i0} = 1)}{d\beta} + (1-p)^2 \frac{dP(\underline{D_i}|G_{i0} = 0)}{d\beta}\right]}{\left(p^2 P(\underline{D_i}|G_{i0} = 2) + 2p(1-p)P(\underline{D_i}|G_{i0} = 1) + (1-p)^2 P(\underline{D_i}|G_{i0} = 0)\right)}$$

The probability of the observed phenotypes in family i, given the proband's genotype, is:

$$P\left(\underline{D_i}\middle|G_{i0}=k\right)=\sum_{\underline{G_i}}P(\underline{G_i}|G_{i0}=k)\prod_{j=0}^{m}P(D_{ij}|G_{ij})$$

Where the "penetrances" are of the form:

$$P\left(D_{ij}\middle|G_{ij}\right)=\exp\left(-\Lambda_0\left(t_{ij}\right)e^{\beta G_{ij}}\right)\left(\lambda_0(t_{ij})e^{\beta G_{ij}}\right)^{D_{ij}}$$

Therefore:

$$P\left(\underline{D_i}\middle|G_{i0}=k\right)=\sum_{\underline{G_i}}P(\underline{G_i}|G_{i0}=k)\prod_{j=0}^{m}\exp\left(-\Lambda_0\left(t_{ij}\right)e^{\beta G_{ij}}\right)\left(\lambda_0(t_{ij})e^{\beta G_{ij}}\right)^{D_{ij}}$$

We note that:

$$\frac{dP(D_{ij}|G_{ij})}{d\beta}=P\left(D_{ij}\middle|G_{ij}\right)G_{ij}\left(D_{ij}-\Lambda_0\left(t_{ij}\right)e^{\beta G_{ij}}\right)$$

So that

$$\begin{aligned}\frac{dP\left(\underline{D_i}\middle|G_{i0}=k\right)}{d\beta}&=\frac{d\left(\sum_{\underline{G_i}}P\left(\underline{G_i}\middle|G_{i0}=k\right)\prod_{j=0}^{m}P\left(D_{ij}\middle|G_{ij}\right)\right)}{d\beta}\\&=\left(\sum_{\underline{G_i}}P\left(\underline{G_i}\middle|G_{i0}=k\right)\frac{d\left(\prod_{j=0}^{m}P\left(D_{ij}\middle|G_{ij}\right)\right)}{d\beta}\right)\sum_{\underline{G_i}}\left(P\left(\underline{G_i}\middle|G_{i0}=k\right)\prod_{j=0}^{m}P\left(D_{ij}\middle|G_{ij}\right)\sum_{j=0}^{m}\frac{1}{P\left(D_{ij}\middle|G_{ij}\right)}\frac{dP\left(D_{ij}\middle|G_{ij}\right)}{d\beta}\right)\\&=\sum_{\underline{G_i}}P\left(\underline{G_i}\middle|G_{i0}=k\right)\left(\prod_{j=0}^{m}P\left(D_{ij}\middle|G_{ij}\right)\sum_{j=0}^{m}G_{ij}\left(D_{ij}-\Lambda_0\left(t_{ij}\right)e^{\beta G_{ij}}\right)\right)\end{aligned}$$

when $\beta\approx0$:

$$\frac{dP\left(\underline{D_i}\middle|G_{i0}=k\right)}{d\beta}=\sum_{\underline{G_i}}P\left(\underline{G_i}\middle|G_{i0}=k\right)\prod_{j=0}^{m}P\left(D_{ij}\right)\sum_{j=0}^{m}G_{ij}\left(D_{ij}-\Lambda_0\left(t_{ij}\right)\right)=\prod_{j=0}^{m}P\left(D_{ij}\right)\sum_{j=0}^{m}\sum_{l=0}^{2}P(G_{ij}=l|G_{i0}=k)G_{ij}\left(D_{ij}-\Lambda_0\left(t_{ij}\right)\right)$$

$$=\prod_{j=0}^{m}P\left(D_{ij}\right)\sum_{j=0}^{m}\left(D_{ij}-\Lambda_0\left(t_{ij}\right)\right)\sum_{l=0}^{2}P(G_{ij}=l|G_{i0}=k)\,l$$

and hence

$$\frac{dlogP\left(\underline{D_i}\middle|G_{i0}=k\right)}{d\beta}=\left.\frac{dP\left(\underline{D_i}\middle|G_{i0}=k\right)}{d\beta}\middle/P\left(\underline{D_i}\middle|G_{i0}=k\right)\right.=\frac{=\prod_{j=0}^{m}P\left(D_{ij}\right)\sum_{j=0}^{m}\left(D_{ij}-\Lambda_0\left(t_{ij}\right)\right)\sum_{l=0}^{2}P(G_{ij}=l|G_{i0}=k)\,l}{\prod_{j=0}^{m}P\left(D_{ij}\right)}=$$

$$=\sum_{j=0}^{m}\left(D_{ij}-\Lambda_0\left(t_{ij}\right)\right)\sum_{l=0}^{2}P(G_{ij}=l|G_{i0}=k)l$$

Therefore:

$$U=\frac{dlogL}{d\beta}_{\beta=0}=\sum_i\frac{1}{P\left(\underline{D_i}\middle|G_{i0}=g_{i0}\right)}\frac{dP\left(\underline{D_i}\middle|G_{i0}=g_{i0}\right)}{d\beta}-\frac{\left[p^2\frac{dP\left(\underline{D_i}\middle|G_{i0}=2\right)}{d\beta}+2p(1-p)\frac{dP\left(\underline{D_i}\middle|G_{i0}=1\right)}{d\beta}+(1-p)^2\frac{dP\left(\underline{D_i}\middle|G_{i0}=0\right)}{d\beta}\right]}{\left(p^2P\left(\underline{D_i}\middle|G_{i0}=2\right)+2p(1-p)P\left(\underline{D_i}\middle|G_{i0}=1\right)+(1-p)^2P\left(\underline{D_i}\middle|G_{i0}=0\right)\right)}=$$

$$=\sum_i\sum_{j=0}^{m}\left(D_{ij}-\Lambda_0\left(t_{ij}\right)\right)\sum_{l=0}^{2}P(G_{ij}=l|G_{i0}=g_{i0})\,l$$

$$-\sum_{j=0}^{m}\left(D_{ij}-\Lambda_0(t_{ik})\right)\left[p^2\sum_{l=0}^{2}P(G_{ij}=l|G_{i0}=2)l+2p(1-p)\sum_{l=0}^{2}P(G_{ij}=l|G_{i0}=1)l+(1-p)^2\sum_{l=0}^{2}P(G_{ij}=l|G_{i0}=0)\,l\right]$$

Since $P\left(\underline{D_i}\middle|G_{i0}\right)=P(\underline{D_i})$ is independent of genotype $G_{i0}$ under the null.

$$\sum_{l=0}^{2} P(G_{ij} = l | G_{i0} = g_{i0})\, l = 2\phi_{i0j} g_{i0} + 2(1 - 2\phi_{i0j})p$$

where $\phi_{i0j}$ is the kinship coefficient between individual j and the proband, and:

$$p^2 \sum_{l=0}^{2} P(G_{ij} = l | G_{i0} = 2) l + 2p(1-p) \sum_{l=0}^{2} P(G_{ij} = l | G_{i0} = 1) l + (1-p)^2 \sum_{l=0}^{2} P(G_{ij} = l | G_{i0} = 0)\, l$$

$$= p^2 (4\phi_{i0j} + 2(1 - 2\phi_{i0j})p) + 2p(1-p) \left(2\phi_{i0j} + 2(1 - 2\phi_{i0j})\right) + (1-p)^2 2(1 - 2\phi_{i0j})p$$
$$= 4p\phi_{i0j} + 2(1 - 2\phi_{i0j})p = 2p$$

Hence:

$$U = \sum_{i} \sum_{j=0}^{m} \left(D_{ij} - \Lambda_0(t_{ij})\right) 2\phi_{i0j}(g_{i0} - 2p)$$

as required.

#### Theoretical power

In the simple case control analysis, excluding family history, the standard test is of the form:

$$U = \sum_i (d_i - \bar{d})(g_i - \bar{g}) = \sum_i d_i' g_i'$$

With variance: $V = Nvar(\underline{d'})var(\underline{g'})$

Where $d_i' = d_i - \bar{d}$ and $g_i' = g_i - \bar{g}$ are normalised phenotypes and genotypes with mean 0, and $N = N_0 + N_1$. This gives a Z-score of the form:

$$Z = U/\sqrt{V}$$

Under the alternative hypothesis, the expected genotype frequency in cases is:

$$2P(g_i = 2|D_i = 1) + P(g_i = 1|D_i = 1) = \frac{2p^2e^{2\beta} + 2p(1-p)e^{\beta}}{p^2e^{2\beta} + 2p(1-p)e^{\beta} + (1-p)^2}$$
$$= \frac{2pe^{\beta}(pe^{\beta} + 1 - p)}{(pe^{\beta} + 1 - p)^2} = \frac{2pe^{\beta}}{pe^{\beta} + 1 - p}$$
$$\sim 2p + 2p(1-p)\beta$$

for small $\beta$.

$E(g_i|d_i) = 2p + 2p(1-p)\beta d_i$ and $E(g_i'|d_i') = 2p(1-p)\beta d_i'$. Therefore:

$$E(Z|\beta) = \frac{EU(\beta)}{\sqrt{V}} = \beta\sqrt{2p(1-p)Nvar(\underline{d'})}$$

The phenotypic variance $var(\underline{d'}) = var(\underline{d}) = \frac{N_1}{N_1+N_0} - \left(\frac{N_1}{N_1+N_0}\right)^2 = \frac{N_0N_1}{N^2}$. Therefore,

$$E(Z|\beta) = \beta\sqrt{2p(1-p)\frac{N_0N_1}{N}}$$

And therefore

$$N_{eff1} = \frac{2N_0N_1}{N_1 + N_0} = 2\left(\frac{1}{N_1} + \frac{1}{N_0}\right)^{-1}$$

Now we consider the test for our models, which is instead based on:

$$U = \sum_i (u_i - \bar{u})(g_i - \bar{g}) = \sum_i u_i' g_i'$$

Where the phenotype $u_i = d_i + \frac{1}{2}f_i$, with family history weighted by ½.

Using the same argument, if $p$ is the allele frequency in controls without a family history, then the expected genotype among controls with a family history is $\sim 2p + p(1-p)\beta$. In controls without family history is $\sim 2p$, in cases without family history is $\sim 2p + 2p(1-p)\beta$ and in cases with a family history is $\sim 2p + 3p(1-p)\beta$.

Therefore, $E(g_i|u_i) = 2p + 2p(1-p)\beta u_i$ and $E(g_i'|u_i') = 2p(1-p)\beta d u_i'$.

This leads, by the same argument as above, to the expected Z-score:

$$E(Z|\beta) = \beta\sqrt{2p(1-p)Nvar(\underline{u'})}$$

$$var(\underline{u'}) = var(\underline{d}) + \frac{1}{4}var\left(\underline{f}\right) + cov\left(\underline{d}, \underline{f}\right)$$
$$= \frac{1}{4N^2}(n_0n_1 + n_1n_2 + n_2n_3 + 4(n_0n_2 + n_1n_3) + 9n_0n_3)$$

Therefore, the effective sample size is:

$$N_{eff2} = 2Nvar(\underline{u'}) = \frac{1}{2N}(n_0n_1 + n_1n_2 + n_2n_3 + 4(n_0n_2 + n_1n_3) + 9n_0n_3)$$

It also possible to also calculate an effective population size if instead, the approach of Liu et al (Liu et al., 2017) of simply grouping cases and controls with a family history ("proxy cases") together, is used. Here the phenotype $y_i = \max(d_i, f_i)$.

In this case the test statistic is of the form:

$$U = \sum_i (y_i - \bar{y})(g_i - \bar{g}) = \sum_i y_i' g_i'$$

With variance:

$$V' = Nvar(y_i)var(g)$$

$E(g_i'|u_i') = 2p(1-p)\beta d u_i'$ and therefore $E(U|\beta) = 2p(1-p)Ncov(\underline{u}, \underline{y})$. So:

$$E(Z|\beta) = cov(\underline{u}, \underline{y})\sqrt{\frac{2p(1-p)N}{var(\underline{y})}}$$

$$cov\left(\underline{u}, \underline{y}\right) = \frac{n_0(n_1 + 2n_2 + 3n_3)}{2N^2}, var\left(\underline{y}\right) = \frac{n_0(n_1 + n_2 + n_3)}{N^2}$$

Therefore, the effective sample size is,

$$N_{eff3} = 2N\frac{cov(\underline{u}, \underline{y})^2}{var(\underline{y})} = \frac{n_0(n_1 + 2n_2 + 3n_3)^2}{2N(n_1 + n_2 + n_3)}$$

Note that Lui *et al* assume in their calculations that using proxy cases will have the same effective sample size as if they were "real" cases, but this overestimates the power since proxy cases will be associated with a smaller effect size. case-control study.

### Supplementary Tables

**Table 1 | ICD-10 codes and UK Biobank self-reported cancer codes used for the cancer definitions.** Self-reported cancer was only included for breast cancer.

| | | |
|---|---|---|
| **Breast cancer** | | C50 |
| | | D05 |
| | | SR1002 |
| **Prostate cancer** | | C61 |
| **Bowel cancer** | Large bowel / colon cancer | C18 |
| | Rectal cancer | C19 |
| | | C20 |
| | Anal cancer | C21 |
| **Lung cancer** | | C34 |

**Table 2 |** Comparing the true effective sample size for the method of Liu et al, with their calculation.

| Cancer | Control | | Case | | $N_{eff3}$ | $N_{eff4}$ |
|---|---|---|---|---|---|---|
| | FH=0 | FH=1 | FH=0 | FH=1 | | |
| **Breast** | 358,515 | 42,740 | 14,800 | 3,252 | 47,396 | 103,956 |
| **Bowel** | 365,391 | 46,969 | 5,761 | 1,186 | 31,113 | 93,967 |
| **Prostate** | 375,319 | 32,323 | 9,945 | 1,720 | 33,490 | 78,747 |
| **Lung** | 361,584 | 54,066 | 2,841 | 816 | 28,895 | 99,553 |

**Table 3 | The power to detect different ORs for breast cancer risk at a significance level of 0.05 using model 3 with varying k values**. Power is calculated by Monte Carlo simulations for 5,000 datasets of size 450,000 with proportions of sex, case/control, and family history the same as for breast cancer in the UK Biobank. For each simulated dataset model 3 was compared to the null model by LRT.

| | | **K=0.3** | **K=0.4** | **K=0.5** | **K=0.6** | **K=0.7** | **K=0.8** | **K=0.9** | **K=1** |
|---|---|---|---|---|---|---|---|---|---|
| **OR** | **1.1** | 0.080 | 0.082 | 0.084 | 0.085 | 0.087 | 0.085 | 0.086 | 0.079 |
| | **1.25** | 0.25 | 0.26 | 0.25 | 0.27 | 0.26 | 0.26 | 0.25 | 0.24 |
| | **1.5** | 0.69 | 0.70 | 0.71 | 0.69 | 0.70 | 0.69 | 0.67 | 0.66 |
| | **1.75** | 0.94 | 0.94 | 0.95 | 0.95 | 0.94 | 0.94 | 0.93 | 0.92 |
| | **2** | 0.99 | 0.99 | 1 | 1 | 0.99 | 0.99 | 0.99 | 0.99 |
| | **2.25** | 1 | 1 | 1 | 1 | 1 | 1 | 1 | 1 |
| | **2.5** | 1 | 1 | 1 | 1 | 1 | 1 | 1 | 1 |

**Table 4 | The power to detect different ORs for breast cancer risk at exome-wide significance, using model 3 with varying k values.** Power is calculated by Monte Carlo simulations for 5,000 datasets of size 450,000 with proportions of sex, case/control, and family history the same as for breast cancer in the UK Biobank. For each simulated dataset model 3 was compared to the null model by LRT.

| | | K=0.3 | K=0.4 | K=0.5 | K=0.6 | K=0.7 | K=0.8 | K=0.9 | K=1 |
|---|---|---|---|---|---|---|---|---|---|
| OR | 1.1 | 0 | 0 | 0 | 0 | 0 | 0 | 0 | 0 |
| | 1.25 | 4.00E-04 | 2.00E-04 | 6.00E-04 | 6.00E-04 | 2.00E-04 | 2.00E-04 | 4.00E-04 | 4.00E-04 |
| | 1.5 | 0.017 | 0.013 | 0.020 | 0.021 | 0.013 | 0.014 | 0.015 | 0.011 |
| | 1.75 | 0.15 | 0.16 | 0.16 | 0.16 | 0.15 | 0.14 | 0.13 | 0.12 |
| | 2 | 0.49 | 0.52 | 0.51 | 0.51 | 0.48 | 0.46 | 0.44 | 0.43 |
| | 2.25 | 0.8 | 0.83 | 0.83 | 0.84 | 0.82 | 0.81 | 0.78 | 0.75 |
| | 2.5 | 0.95 | 0.97 | 0.97 | 0.97 | 0.96 | 0.96 | 0.94 | 0.94 |

**Table 5 | Association results for PTVs and breast cancer using model 3 with k=0.5 and model 1.** All genes associated at P<0.001 are listed. Z-scores and p-values are from testing H0:β=ln(Odds Ratio)=0 (2-tailed). All p-values are unadjusted for multiple testing. The counts are from female cases and controls, but results are from incorporating males and family history data. The table is sorted by ascending P-value for model 3.

| Gene | Controls | | Cases | | Model 1 | | Model 3 | |
|---|---|---|---|---|---|---|---|---|
| | Non-Carriers | Carriers | Non-Carriers | Carriers | OR | P-value | OR | P-value |
| ***BRCA2*** | 208996 | 439 | 17736 | 222 | 6.12 (5.22, 7.19) | 1.84E-108 | 6.79 (5.98, 7.72) | 3.43E-189 |
| ***BRCA1*** | 209338 | 97 | 17881 | 77 | 9.09 (6.75, 12.2) | 4.98E-48 | 11.3 (8.97, 14.3) | 2.17E-92 |
| ***PALB2*** | 209143 | 292 | 17857 | 101 | 4.03 (3.21, 5.04) | 1.07E-33 | 4.33 (3.62, 5.18) | 6.86E-58 |
| ***CHEK2*** | 208444 | 991 | 17749 | 209 | 2.45 (2.11, 2.84) | 9.99E-32 | 2.49 (2.22, 2.81) | 1.51E-51 |
| ***ATM*** | 208955 | 480 | 17866 | 92 | 2.23 (1.79, 2.79) | 1.74E-12 | 2.32 (1.94, 2.76) | 1.71E-20 |
| ***MAP3K1*** | 209413 | 22 | 17949 | 9 | 4.74 (2.18, 10.3) | 8.30E-05 | 5.63 (3.05, 10.4) | 3.20E-08 |
| ***BAP1*** | 209421 | 14 | 17952 | 6 | 4.97 (1.92, 12.9) | 0.000981 | 5.17 (2.42, 11.1) | 2.30E-05 |
| ***COL12A1*** | 209382 | 53 | 17945 | 13 | 2.83 (1.54, 5.19) | 0.000763 | 2.62 (1.6, 4.3) | 0.000135 |
| ***RNF112*** | 209398 | 37 | 17948 | 10 | 3.07 (1.53, 6.17) | 0.00161 | 2.94 (1.68, 5.13) | 0.000146 |
| ***KCND2*** | 209432 | 3 | 17955 | 3 | 11.8 (2.41, 57.7) | 0.00233 | 11.6 (3.2, 41.9) | 0.000189 |
| ***BARD1*** | 209363 | 72 | 17941 | 17 | 2.72 (1.61, 4.62) | 0.000201 | 2.3 (1.48, 3.58) | 0.000213 |
| ***MMP26*** | 209426 | 9 | 17952 | 6 | 7.97 (2.85, 22.3) | 7.78E-05 | 5.25 (2.16, 12.8) | 0.000262 |
| ***GPR37*** | 209429 | 6 | 17955 | 3 | 5.65 (1.42, 22.5) | 0.0142 | 7.25 (2.46, 21.4) | 0.000333 |
| ***CYBC1*** | 209433 | 2 | 17956 | 2 | 11.8 (1.66, 83.3) | 0.0137 | 25.3 (4.31, 148) | 0.000346 |
| ***SEC62*** | 209433 | 2 | 17956 | 2 | 11.4 (1.62, 80.3) | 0.0146 | 20.8 (3.89, 111) | 0.000384 |
| ***PCDHGB3*** | 208970 | 465 | 17903 | 55 | 1.36 (1.02, 1.8) | 0.0329 | 1.47 (1.19, 1.82) | 0.000422 |
| ***CFAP126*** | 209416 | 19 | 17951 | 7 | 4.34 (1.83, 10.3) | 0.000877 | 3.7 (1.78, 7.71) | 0.000470 |
| ***FLYWCH2*** | 209430 | 5 | 17955 | 3 | 12.2 (3.06, 48.3) | 0.00039 | 7.83 (2.42, 25.4) | 0.000604 |
| ***KLK4*** | 209360 | 75 | 17945 | 13 | 1.99 (1.1, 3.58) | 0.0219 | 2.19 (1.4, 3.42) | 0.000615 |
| ***LSP1*** | 209370 | 65 | 17947 | 11 | 2 (1.06, 3.79) | 0.033 | 2.3 (1.42, 3.71) | 0.000667 |
| ***CHTF18*** | 209098 | 337 | 17945 | 13 | 0.446 (0.256, 0.776) | 0.00425 | 0.515 (0.352, 0.756) | 0.000689 |
| ***FNDC3A*** | 209432 | 3 | 17954 | 4 | 15 (3.42, 65.5) | 0.000328 | 8.01 (2.4, 26.7) | 0.000723 |
| ***MGAT5*** | 209421 | 14 | 17955 | 3 | 2.51 (0.72, 8.73) | 0.149 | 4.46 (1.86, 10.6) | 0.000772 |
| ***KRT28*** | 209244 | 191 | 17950 | 8 | 0.48 (0.237, 0.975) | 0.0422 | 0.391 (0.226, 0.677) | 0.000789 |
| ***NOTCH3*** | 209387 | 48 | 17945 | 13 | 3.13 (1.7, 5.77) | 0.000262 | 2.48 (1.46, 4.22) | 0.000793 |
| ***CDH1*** | 209427 | 8 | 17954 | 4 | 5.9 (1.79, 19.5) | 0.00358 | 5.14 (1.97, 13.4) | 0.000837 |
| ***CDX1*** | 209429 | 6 | 17955 | 3 | 5.66 (1.42, 22.6) | 0.0139 | 6.36 (2.12, 19.1) | 0.000983 |
| ***STPG4*** | 209346 | 89 | 17940 | 18 | 2.38 (1.43, 3.94) | 0.000808 | 1.98 (1.28, 3.04) | 0.00198 |
| ***UBE2Q2*** | 209399 | 36 | 17947 | 11 | 3.53 (1.8, 6.93) | 0.000247 | 2.54 (1.39, 4.64) | 0.00240 |
| ***FAM9A*** | 209252 | 183 | 17928 | 30 | 1.92 (1.31, 2.83) | 0.000914 | 1.68 (1.19, 2.38) | 0.00296 |
| ***DRG2*** | 209419 | 16 | 17951 | 7 | 5.17 (2.13, 12.5) | 0.000282 | 3.26 (1.46, 7.27) | 0.00392 |
| ***MORC1*** | 209334 | 101 | 17935 | 23 | 2.61 (1.66, 4.11) | 3.21E-05 | 1.77 (1.17, 2.66) | 0.00639 |
| ***ZC2HC1B*** | 209401 | 34 | 17948 | 10 | 3.42 (1.69, 6.92) | 0.00062 | 2.32 (1.22, 4.41) | 0.0103 |
| ***ENTPD5*** | 209359 | 76 | 17941 | 17 | 2.56 (1.51, 4.33) | 0.000454 | 1.79 (1.12, 2.84) | 0.0143 |
| ***ANKRD30A*** | 188474 | 20961 | 16309 | 1649 | 0.912 (0.865, 0.961) | 0.000564 | 0.955 (0.918, 0.993) | 0.0214 |
| ***NPSR1*** | 209295 | 140 | 17933 | 25 | 2.05 (1.34, 3.14) | 0.000933 | 1.35 (0.917, 1.99) | 0.128 |

**Table 6 | Association results for PTVs and prostate cancer using model 3 with k=0.5 and model 1.** All genes associated at P<0.001 are listed. Z-scores and p-values are from testing H0:β=ln(Odds Ratio)=0 (2-tailed). All p-values are unadjusted for multiple testing. The counts are from male cases and controls, but results are from incorporating females and family history data. The table is sorted by ascending P-value for model 3.

| Gene | Controls | | Cases | | Model 1 | | Model 3 | |
|---|---|---|---|---|---|---|---|---|
| | Non-Carriers | Carriers | Non-Carriers | Carriers | OR | P-value | OR | P-value |
| ***BRCA2*** | 179731 | 518 | 11591 | 74 | 2.22 (1.74, 2.84) | 1.54E-10 | 2.27 (1.88, 2.75) | 2.50E-17 |
| ***CHEK2*** | 179287 | 962 | 11546 | 119 | 1.88 (1.55, 2.27) | 1.26E-10 | 1.65 (1.41, 1.93) | 3.59E-10 |
| ***ATM*** | 179859 | 390 | 11618 | 47 | 1.87 (1.38, 2.54) | 4.91E-05 | 1.93 (1.53, 2.43) | 2.26E-08 |
| ***GEMIN2*** | 180238 | 11 | 11660 | 5 | 7 (2.43, 20.2) | 0.000313 | 7.21 (3.12, 16.7) | 3.92E-06 |
| ***OSGIN1*** | 180238 | 11 | 11661 | 4 | 5.69 (1.81, 17.9) | 0.00292 | 6.5 (2.74, 15.4) | 2.11E-05 |
| ***UBQLN4*** | 180247 | 2 | 11661 | 4 | 29.3 (5.35, 161) | 9.91E-05 | 14.7 (4.05, 53.3) | 4.34E-05 |
| ***C9orf50*** | 180228 | 21 | 11660 | 5 | 3.83 (1.44, 10.2) | 0.00704 | 4.31 (2.1, 8.87) | 7.12E-05 |
| ***C9orf152*** | 173671 | 6578 | 11197 | 468 | 1.09 (0.99, 1.2) | 0.0788 | 1.15 (1.07, 1.24) | 0.000101 |
| ***SNX2*** | 180237 | 12 | 11662 | 3 | 3.89 (1.1, 13.8) | 0.0355 | 5.91 (2.4, 14.6) | 0.000112 |
| ***MFSD8*** | 180219 | 30 | 11657 | 8 | 4.04 (1.85, 8.81) | 0.000459 | 3.46 (1.81, 6.61) | 0.000169 |
| ***GRIFIN*** | 180247 | 2 | 11663 | 2 | 16.8 (2.34, 120) | 0.00498 | 18.4 (3.81, 89.3) | 0.000291 |
| ***NOCT*** | 180231 | 18 | 11661 | 4 | 3.35 (1.13, 9.91) | 0.0288 | 4.22 (1.92, 9.29) | 0.000339 |
| ***PPP5C*** | 180105 | 144 | 11649 | 16 | 1.71 (1.01, 2.89) | 0.047 | 2.02 (1.37, 2.98) | 0.000352 |
| ***INVS*** | 180070 | 179 | 11641 | 24 | 2.08 (1.36, 3.19) | 0.000754 | 1.87 (1.32, 2.64) | 0.000363 |
| ***DEGS1*** | 180235 | 14 | 11661 | 4 | 4.31 (1.42, 13.1) | 0.00998 | 4.89 (2.04, 11.7) | 0.000379 |
| ***PTS*** | 180244 | 5 | 11662 | 3 | 9.76 (2.31, 41.2) | 0.00193 | 8.85 (2.66, 29.5) | 0.000385 |
| ***TNFRSF11A*** | 180239 | 10 | 11662 | 3 | 4.76 (1.31, 17.3) | 0.018 | 5.98 (2.22, 16.1) | 0.000400 |
| ***NUP98*** | 180245 | 4 | 11662 | 3 | 11.4 (2.55, 51) | 0.00146 | 9.18 (2.69, 31.3) | 0.000403 |
| ***MALL*** | 180247 | 2 | 11662 | 3 | 23.6 (3.9, 142) | 0.000578 | 14.7 (3.29, 65.8) | 0.000431 |
| ***MYH7*** | 180145 | 104 | 11648 | 17 | 2.53 (1.52, 4.23) | 0.000386 | 2.16 (1.4, 3.33) | 0.000459 |
| ***GOLPH3L*** | 180228 | 21 | 11660 | 5 | 3.51 (1.32, 9.32) | 0.0116 | 3.92 (1.82, 8.45) | 0.000489 |
| ***C20orf27*** | 180247 | 2 | 11663 | 2 | 16.1 (2.27, 115) | 0.00547 | 14.1 (3.14, 63.7) | 0.000563 |
| ***ESCO1*** | 180236 | 13 | 11660 | 5 | 5.96 (2.12, 16.7) | 0.000704 | 4.49 (1.9, 10.6) | 0.000626 |
| ***SMAD2*** | 180247 | 2 | 11662 | 3 | 23.4 (3.9, 141) | 0.000564 | 12.4 (2.92, 53) | 0.000645 |
| ***PNLDC1*** | 180124 | 125 | 11647 | 18 | 2.17 (1.32, 3.55) | 0.00218 | 2 (1.34, 2.97) | 0.000657 |
| ***CMTM2*** | 180216 | 33 | 11660 | 5 | 2.37 (0.923, 6.06) | 0.0729 | 3.19 (1.63, 6.21) | 0.000676 |
| ***BET1*** | 180129 | 120 | 11650 | 15 | 1.9 (1.11, 3.25) | 0.0194 | 2.03 (1.35, 3.06) | 0.000684 |
| ***PDHB*** | 180241 | 8 | 11663 | 2 | 3.87 (0.821, 18.2) | 0.0871 | 6.27 (2.16, 18.2) | 0.000751 |
| ***GFRA1*** | 180244 | 5 | 11663 | 2 | 6.12 (1.19, 31.6) | 0.0304 | 7.4 (2.3, 23.8) | 0.000781 |
| ***MBP*** | 180246 | 3 | 11662 | 3 | 15.2 (3.07, 75.4) | 0.000863 | 8.49 (2.44, 29.6) | 0.000785 |
| ***B4GALT6*** | 180235 | 14 | 11661 | 4 | 4.3 (1.42, 13.1) | 0.0101 | 4.61 (1.88, 11.3) | 0.000838 |
| ***HSPA12B*** | 180238 | 11 | 11660 | 5 | 7.34 (2.55, 21.1) | 0.000223 | 4.72 (1.9, 11.8) | 0.000861 |
| ***BSCL2*** | 180212 | 37 | 11661 | 4 | 1.67 (0.593, 4.68) | 0.333 | 2.96 (1.56, 5.62) | 0.000882 |
| ***EMP3*** | 180226 | 23 | 11660 | 5 | 3.26 (1.24, 8.6) | 0.0166 | 3.51 (1.67, 7.39) | 0.000926 |
| ***FOXR1*** | 180199 | 50 | 11654 | 11 | 3.33 (1.73, 6.4) | 0.000304 | 2.46 (1.42, 4.26) | 0.00134 |
| ***HEATR3*** | 180220 | 29 | 11657 | 8 | 4.18 (1.91, 9.15) | 0.000344 | 2.99 (1.52, 5.88) | 0.00144 |
| ***MICB*** | 180031 | 218 | 11635 | 30 | 2.14 (1.46, 3.14) | 9.61E-05 | 1.68 (1.22, 2.33) | 0.00160 |
| ***SUN5*** | 180214 | 35 | 11656 | 9 | 3.92 (1.88, 8.15) | 0.000264 | 2.83 (1.48, 5.42) | 0.00170 |
| ***GPR153*** | 180236 | 13 | 11660 | 5 | 6.06 (2.16, 17) | 0.000623 | 3.83 (1.54, 9.49) | 0.00378 |
| ***MIB2*** | 180205 | 44 | 11654 | 11 | 3.8 (1.96, 7.37) | 7.59E-05 | 2.29 (1.23, 4.26) | 0.00929 |
| ***RERE*** | 180196 | 53 | 11654 | 11 | 3.29 (1.72, 6.3) | 0.000329 | 2.13 (1.18, 3.84) | 0.0118 |
| ***RUFY1*** | 180072 | 177 | 11641 | 24 | 2.08 (1.36, 3.19) | 0.000758 | 1.58 (1.1, 2.28) | 0.0128 |
| ***POLR3B*** | 180182 | 67 | 11653 | 12 | 2.81 (1.52, 5.2) | 0.000974 | 1.92 (1.11, 3.31) | 0.0199 |
| ***FBXO17*** | 180152 | 97 | 11650 | 15 | 2.52 (1.46, 4.34) | 0.000888 | 1.75 (1.09, 2.82) | 0.0215 |
| ***HMX3*** | 180235 | 14 | 11659 | 6 | 6.52 (2.5, 17) | 0.000123 | 2.85 (1.12, 7.25) | 0.0283 |
| ***UNC5CL*** | 180034 | 215 | 11637 | 28 | 1.95 (1.31, 2.89) | 0.000916 | 1.34 (0.941, 1.9) | 0.105 |

**Table 7 | Association results for PTVs and lung cancer using model 3 with k=0.5 and model 1.** All genes associated at P<0.001 for model 3 or model 1 are listed. Z-scores and p-values are from testing H0:β=ln(Odds Ratio)=0 (2-tailed). All p-values are unadjusted for multiple testing. The counts and results are from data for males and females. The table is sorted by ascending P-value for model 3.

| Gene | Controls | | Cases | | Model 1 | | Model 3 | |
|---|---|---|---|---|---|---|---|---|
| | Non-Carriers | Carriers | Non-Carriers | Carriers | OR | P-value | OR | P-value |
| ***MON2*** | 415500 | 150 | 3653 | 4 | 3.08 (1.14, 8.31) | 0.0267 | 3.5 (1.98, 6.2) | 1.67E-05 |
| ***ASB6*** | 415599 | 51 | 3655 | 2 | 4.64 (1.13, 19.1) | 0.0334 | 6.05 (2.65, 13.8) | 1.89E-05 |
| ***ABCF2*** | 415623 | 27 | 3655 | 2 | 8.32 (1.98, 35.1) | 0.00389 | 7.97 (2.91, 21.8) | 5.38E-05 |
| ***ARHGAP35*** | 415617 | 33 | 3656 | 1 | 3.65 (0.498, 26.7) | 0.203 | 7.28 (2.74, 19.3) | 6.79E-05 |
| ***KCNH8*** | 415516 | 134 | 3656 | 1 | 0.858 (0.12, 6.14) | 0.879 | 3.45 (1.87, 6.35) | 7.17E-05 |
| ***BIRC3*** | 415625 | 25 | 3655 | 2 | 9.76 (2.31, 41.3) | 0.00196 | 8.36 (2.92, 23.9) | 7.59E-05 |
| ***MXRA5*** | 225605 | 36 | 1751 | 1 | 3.44 (0.472, 25.2) | 0.223 | 6.74 (2.58, 17.6) | 9.78E-05 |
| ***CLU*** | 415614 | 36 | 3655 | 2 | 6.64 (1.6, 27.6) | 0.00925 | 6.56 (2.54, 17) | 0.000102 |
| ***TBRG4*** | 415556 | 94 | 3654 | 3 | 3.74 (1.18, 11.8) | 0.0248 | 3.94 (1.97, 7.9) | 0.000108 |
| ***ATM*** | 414659 | 991 | 3639 | 18 | 2.09 (1.31, 3.34) | 0.00198 | 1.71 (1.3, 2.25) | 0.000124 |
| ***HIVEP3*** | 415546 | 104 | 3652 | 5 | 5.44 (2.21, 13.4) | 0.000231 | 3.66 (1.88, 7.14) | 0.000137 |
| ***SMOX*** | 415602 | 48 | 3655 | 2 | 4.72 (1.14, 19.4) | 0.0318 | 5.42 (2.27, 12.9) | 0.000141 |
| ***KCNIP1*** | 415637 | 13 | 3655 | 2 | 19.8 (4.46, 88.4) | 8.85E-05 | 12 (3.33, 43.4) | 0.000145 |
| ***MAGI1*** | 415606 | 44 | 3654 | 3 | 8.35 (2.59, 26.9) | 0.000384 | 5.7 (2.32, 14) | 0.000149 |
| ***ZGRF1*** | 414047 | 1603 | 3647 | 10 | 0.709 (0.38, 1.32) | 0.279 | 0.57 (0.426, 0.762) | 0.000154 |
| ***NT5C1A*** | 415619 | 31 | 3655 | 2 | 7.12 (1.7, 29.8) | 0.00723 | 6.63 (2.46, 17.8) | 0.000183 |
| ***NUP160*** | 415476 | 174 | 3653 | 4 | 2.61 (0.968, 7.04) | 0.0578 | 2.88 (1.64, 5.06) | 0.000219 |
| ***TRIM47*** | 415545 | 105 | 3651 | 6 | 6.59 (2.89, 15) | 7.38E-06 | 3.53 (1.8, 6.91) | 0.000232 |
| ***ZSCAN26*** | 415615 | 35 | 3655 | 2 | 6.72 (1.61, 28) | 0.00888 | 6.26 (2.36, 16.6) | 0.000236 |
| ***MET*** | 415613 | 37 | 3655 | 2 | 6.11 (1.47, 25.4) | 0.0128 | 5.96 (2.3, 15.4) | 0.000239 |
| ***CLCF1*** | 415640 | 10 | 3655 | 2 | 23.2 (5.05, 106) | 5.25E-05 | 12.4 (3.18, 48.7) | 0.000296 |
| ***RANGRF*** | 411695 | 3955 | 3609 | 48 | 1.38 (1.04, 1.84) | 0.0262 | 1.32 (1.13, 1.53) | 0.000296 |
| ***ADAP1*** | 415619 | 31 | 3655 | 2 | 7.42 (1.77, 31.1) | 0.00607 | 6.47 (2.34, 17.9) | 0.000316 |
| ***TRIM43*** | 415625 | 25 | 3654 | 3 | 13.1 (3.94, 43.6) | 2.70E-05 | 6.97 (2.41, 20.1) | 0.000333 |
| ***DGKI*** | 415576 | 74 | 3654 | 3 | 4.64 (1.46, 14.7) | 0.00923 | 4.03 (1.88, 8.66) | 0.000357 |
| ***SYT1*** | 415649 | 1 | 3656 | 1 | 143 (8.18, 2500) | 0.000676 | 85.9 (7.45, 991) | 0.000358 |
| ***REV3L*** | 415579 | 71 | 3652 | 5 | 7.51 (3.02, 18.6) | 1.39E-05 | 3.92 (1.83, 8.38) | 0.000434 |
| ***CYB5B*** | 415633 | 17 | 3656 | 1 | 6.44 (0.854, 48.5) | 0.0708 | 8.94 (2.64, 30.3) | 0.000435 |
| ***ADGRE2*** | 412089 | 3561 | 3616 | 41 | 1.32 (0.969, 1.8) | 0.0783 | 1.32 (1.13, 1.55) | 0.000469 |
| ***KPNA5*** | 415585 | 65 | 3655 | 2 | 3.51 (0.858, 14.4) | 0.0806 | 4.2 (1.86, 9.5) | 0.000560 |
| ***STAR*** | 415621 | 29 | 3656 | 1 | 3.69 (0.501, 27.1) | 0.2 | 6.32 (2.21, 18) | 0.000578 |
| ***DHH*** | 415646 | 4 | 3655 | 2 | 49.6 (8.93, 276) | 8.18E-06 | 18 (3.43, 94) | 0.000628 |
| ***DEPDC1*** | 415441 | 209 | 3653 | 4 | 2.15 (0.799, 5.79) | 0.13 | 2.53 (1.48, 4.3) | 0.000632 |
| ***AMPD2*** | 415462 | 188 | 3652 | 5 | 3.02 (1.24, 7.34) | 0.0149 | 2.61 (1.5, 4.52) | 0.000635 |
| ***RGS14*** | 415600 | 50 | 3653 | 4 | 9.02 (3.25, 25) | 2.38E-05 | 4.6 (1.91, 11.1) | 0.000653 |
| ***DPY19L4*** | 414906 | 744 | 3646 | 11 | 1.7 (0.935, 3.08) | 0.0819 | 1.74 (1.26, 2.38) | 0.000665 |
| ***ZNF718*** | 415649 | 1 | 3656 | 1 | 118 (7.18, 1940) | 0.000837 | 68.1 (5.96, 778) | 0.000681 |
| ***SPDYE5*** | 415471 | 179 | 3651 | 6 | 3.89 (1.72, 8.79) | 0.00108 | 2.62 (1.49, 4.62) | 0.000840 |
| ***RBM4B*** | 415605 | 45 | 3655 | 2 | 5 (1.21, 20.7) | 0.026 | 4.85 (1.92, 12.2) | 0.000844 |
| ***ATP5F1E*** | 415629 | 21 | 3656 | 1 | 6.2 (0.832, 46.2) | 0.0749 | 7.93 (2.35, 26.8) | 0.000855 |
| ***HDAC5*** | 415637 | 13 | 3656 | 1 | 9.01 (1.17, 69.3) | 0.0346 | 10.2 (2.6, 39.7) | 0.000859 |
| ***SERBP1*** | 415649 | 1 | 3656 | 1 | 323 (9.17, 11400) | 0.00147 | 161 (7.84, 3310) | 0.000982 |
| ***GLRA2*** | 225639 | 2 | 1751 | 1 | 69 (6.02, 792) | 0.000668 | 40.9 (4.49, 373) | 0.000997 |
| ***RNF103*** | 415645 | 5 | 3655 | 2 | 39.7 (7.59, 207) | 1.29E-05 | 15.1 (2.96, 76.9) | 0.00109 |
| ***WNT4*** | 415644 | 6 | 3655 | 2 | 41 (8.22, 205) | 5.98E-06 | 14.8 (2.93, 74.4) | 0.00110 |
| ***TAC1*** | 415641 | 9 | 3655 | 2 | 26.9 (5.78, 125) | 2.72E-05 | 11.6 (2.64, 50.5) | 0.00115 |

| | | | | | | | | |
|---|---|---|---|---|---|---|---|---|
| ***COPS5*** | 415648 | 2 | 3656 | 1 | 69.3 (5.85, 822) | 0.00078 | 40.2 (4.28, 377) | 0.00122 |
| ***MAP2K3*** | 252905 | 162745 | 2120 | 1537 | 1.12 (1.05, 1.2) | 0.000505 | 1.05 (1.02, 1.09) | 0.00123 |
| ***YBEY*** | 415620 | 30 | 3653 | 4 | 16.4 (5.76, 46.8) | 1.64E-07 | 5.75 (1.98, 16.7) | 0.00130 |
| ***DHRS7C*** | 415567 | 83 | 3653 | 4 | 5.57 (2.04, 15.2) | 0.000811 | 3.47 (1.62, 7.44) | 0.00141 |
| ***MAP2K6*** | 415648 | 2 | 3656 | 1 | 59.6 (5.35, 664) | 0.000889 | 32.2 (3.79, 275) | 0.00148 |
| ***MAGI2*** | 415627 | 23 | 3654 | 3 | 15.6 (4.69, 52.3) | 7.78E-06 | 6.29 (1.97, 20.1) | 0.00192 |
| ***SPANXB1*** | 225638 | 3 | 1751 | 1 | 53.5 (5.38, 531) | 0.000682 | 25.4 (2.96, 219) | 0.00321 |
| ***SERPINB8*** | 415540 | 110 | 3652 | 5 | 5.14 (2.09, 12.6) | 0.000353 | 2.86 (1.42, 5.75) | 0.00328 |
| ***EXOC6B*** | 415610 | 40 | 3654 | 3 | 8.52 (2.63, 27.6) | 0.000355 | 4.42 (1.63, 12) | 0.00352 |
| ***DMXL1*** | 415429 | 221 | 3649 | 8 | 4.12 (2.03, 8.35) | 8.68E-05 | 2.17 (1.26, 3.72) | 0.00489 |
| ***CASTOR1*** | 415632 | 18 | 3654 | 3 | 17.2 (5.05, 58.8) | 5.40E-06 | 5.49 (1.5, 20.2) | 0.0102 |
| ***HLCS*** | 415193 | 457 | 3645 | 12 | 3 (1.69, 5.32) | 0.000182 | 1.7 (1.13, 2.54) | 0.0106 |
| ***MICB*** | 415114 | 536 | 3645 | 12 | 2.65 (1.49, 4.7) | 0.000884 | 1.65 (1.12, 2.42) | 0.0109 |
| ***PDGFC*** | 415642 | 8 | 3655 | 2 | 25.7 (5.43, 121) | 4.25E-05 | 8.14 (1.57, 42.2) | 0.0125 |
| ***CDK14*** | 415603 | 47 | 3654 | 3 | 7.5 (2.33, 24.1) | 0.00073 | 3.55 (1.3, 9.69) | 0.0133 |
| ***ACTN1*** | 415634 | 16 | 3655 | 2 | 14.3 (3.27, 62.4) | 0.000404 | 5.86 (1.44, 23.8) | 0.0133 |
| ***MGLL*** | 415641 | 9 | 3655 | 2 | 24.9 (5.34, 116) | 4.21E-05 | 7.91 (1.53, 40.8) | 0.0135 |
| ***SIX4*** | 415626 | 24 | 3654 | 3 | 13.6 (4.07, 45.2) | 2.15E-05 | 4.54 (1.32, 15.6) | 0.0163 |
| ***GALR3*** | 415643 | 7 | 3655 | 2 | 39.1 (7.98, 191) | 6.11E-06 | 9.08 (1.44, 57.1) | 0.0187 |
| ***DNAJC5G*** | 415632 | 18 | 3655 | 2 | 12.5 (2.88, 53.9) | 0.000732 | 4.99 (1.23, 20.3) | 0.0247 |
| ***EIF1B*** | 415648 | 2 | 3656 | 1 | 66.2 (5.74, 764) | 0.00078 | 17.6 (1.38, 224) | 0.0272 |
| ***NFKBIB*** | 415633 | 17 | 3655 | 2 | 12.2 (2.8, 53) | 0.00086 | 4.83 (1.19, 19.6) | 0.0276 |
| ***OTUD5*** | 225639 | 2 | 1751 | 1 | 68.1 (6.09, 762) | 0.000613 | 17.4 (1.32, 230) | 0.0299 |
| ***STAG1*** | 415648 | 2 | 3656 | 1 | 60.6 (5.37, 684) | 0.000901 | 16.6 (1.31, 210) | 0.0300 |
| ***CAMLG*** | 415583 | 67 | 3653 | 4 | 6.11 (2.22, 16.8) | 0.000451 | 2.62 (1.09, 6.27) | 0.0308 |
| ***IRF2*** | 415648 | 2 | 3656 | 1 | 63.8 (5.6, 726) | 0.000813 | 15.8 (1.24, 200) | 0.0333 |
| ***SIPA1L2*** | 415610 | 40 | 3654 | 3 | 8.43 (2.6, 27.3) | 0.000381 | 3.2 (1.07, 9.54) | 0.0373 |
| ***ZNF106*** | 415570 | 80 | 3653 | 4 | 5.49 (2.01, 15) | 0.000908 | 2.44 (1.04, 5.7) | 0.0400 |
| ***KAT2B*** | 415579 | 71 | 3653 | 4 | 6.42 (2.34, 17.6) | 0.000304 | 2.52 (1.03, 6.17) | 0.0425 |
| ***TIMM10B*** | 415617 | 33 | 3654 | 3 | 10.7 (3.28, 35.1) | 8.55E-05 | 3.42 (1.04, 11.2) | 0.0431 |
| ***SEC24B*** | 415606 | 44 | 3654 | 3 | 7.85 (2.43, 25.3) | 0.000564 | 2.94 (0.988, 8.72) | 0.0525 |
| ***HOMER2*** | 415564 | 86 | 3653 | 4 | 5.49 (2.01, 15) | 0.000881 | 2.3 (0.983, 5.39) | 0.0549 |
| ***DDR2*** | 415614 | 36 | 3654 | 3 | 9.91 (3.05, 32.3) | 0.000139 | 3.17 (0.968, 10.4) | 0.0567 |
| ***TRMT2B*** | 225620 | 21 | 1750 | 2 | 12.1 (2.84, 51.9) | 0.000761 | 3.98 (0.952, 16.7) | 0.0585 |
| ***SERPINB6*** | 415632 | 18 | 3655 | 2 | 13.1 (3.02, 56.6) | 0.000578 | 4.21 (0.926, 19.1) | 0.0628 |
| ***ZNF75D*** | 225618 | 23 | 1750 | 2 | 11.7 (2.75, 49.9) | 0.000875 | 3.77 (0.902, 15.8) | 0.0689 |
| ***GBF1*** | 415482 | 168 | 3651 | 6 | 4.18 (1.85, 9.44) | 0.000591 | 1.71 (0.876, 3.33) | 0.116 |
| ***SYNGR3*** | 415632 | 18 | 3655 | 2 | 14.2 (3.29, 61.6) | 0.000379 | 3.16 (0.582, 17.1) | 0.183 |
| ***GP2*** | 415531 | 119 | 3652 | 5 | 5.03 (2.05, 12.3) | 0.00041 | 1.65 (0.741, 3.68) | 0.220 |
| ***ATF6B*** | 415532 | 118 | 3652 | 5 | 4.64 (1.89, 11.4) | 0.000788 | 1.46 (0.654, 3.27) | 0.354 |
| ***GALNT7*** | 415571 | 79 | 3653 | 4 | 5.84 (2.13, 16) | 0.000587 | 1.34 (0.478, 3.75) | 0.580 |

**Table 8 | Association results for PTVs and bowel cancer using model 3 with k=0.5 and model 1.** All genes associated at P<0.001 are listed. Z-scores and p-values are from testing H0:β=ln(Odds Ratio)=0 (2-tailed). All p-values are unadjusted for multiple testing. The counts and results are from data for males and females. The table is sorted by ascending P-value for model 3.

| Gene | Controls | | Cases | | Model 1 | | Model 3 | |
|---|---|---|---|---|---|---|---|---|
| | Non-Carriers | Carriers | Non-Carriers | Carriers | OR | P-value | OR | P-value |
| ***MSH6*** | 412066 | 294 | 6913 | 34 | 6.94 (4.86, 9.91) | 1.92E-26 | 7.47 (5.71, 9.78) | 1.76E-48 |
| ***MSH2*** | 412230 | 130 | 6928 | 19 | 8.5 (5.24, 13.8) | 4.32E-18 | 9.11 (6.29, 13.2) | 1.79E-31 |
| ***MLH1*** | 412349 | 11 | 6938 | 9 | 50.1 (20.6, 122) | 5.78E-18 | 61.7 (28.7, 133) | 5.56E-26 |
| ***APC*** | 412335 | 25 | 6937 | 10 | 25.2 (12, 52.7) | 1.18E-17 | 20.8 (11, 39.2) | 5.18E-21 |
| ***GAPDH*** | 412281 | 79 | 6940 | 7 | 5.43 (2.5, 11.8) | 1.90E-05 | 4.67 (2.52, 8.64) | 9.29E-07 |
| ***MT1G*** | 412358 | 2 | 6945 | 2 | 54.8 (7.56, 397) | 7.40E-05 | 43.3 (8.02, 234) | 1.20E-05 |
| ***FLCN*** | 412250 | 110 | 6939 | 8 | 4.18 (2.03, 8.58) | 9.87E-05 | 3.51 (1.99, 6.19) | 1.42E-05 |
| ***SMAD4*** | 412356 | 4 | 6945 | 2 | 30 (5.42, 166) | 9.90E-05 | 22 (4.94, 98.2) | 5.02E-05 |
| ***ATF3*** | 412342 | 18 | 6945 | 2 | 6.15 (1.42, 26.6) | 0.015 | 7.9 (2.8, 22.3) | 9.16E-05 |
| ***CNTFR*** | 412348 | 12 | 6944 | 3 | 15.4 (4.3, 54.8) | 2.60E-05 | 9.87 (3.1, 31.4) | 0.00010 |
| ***CTTNBP2NL*** | 412356 | 4 | 6946 | 1 | 16.5 (1.81, 150) | 0.0129 | 24.3 (4.68, 126) | 0.000146 |
| ***NPNT*** | 412292 | 68 | 6944 | 3 | 2.62 (0.824, 8.35) | 0.103 | 3.95 (1.94, 8.04) | 0.000152 |
| ***FRRS1*** | 412290 | 70 | 6942 | 5 | 4.32 (1.74, 10.7) | 0.0016 | 3.86 (1.92, 7.78) | 0.000156 |
| ***CLEC4A*** | 412305 | 55 | 6942 | 5 | 5.48 (2.19, 13.7) | 0.000282 | 4.25 (1.99, 9.08) | 0.000182 |
| ***PMS2*** | 412147 | 213 | 6934 | 13 | 3.59 (2.05, 6.29) | 8.25E-06 | 2.44 (1.52, 3.89) | 0.000195 |
| ***BARD1*** | 412192 | 168 | 6937 | 10 | 3.48 (1.84, 6.6) | 0.000132 | 2.64 (1.58, 4.42) | 0.000212 |
| ***SNRPC*** | 412356 | 4 | 6946 | 1 | 14.4 (1.59, 130) | 0.0177 | 21.4 (4.2, 110) | 0.000229 |
| ***RALGPS2*** | 412317 | 43 | 6944 | 3 | 4.53 (1.4, 14.6) | 0.0116 | 4.83 (2.08, 11.2) | 0.000259 |
| ***C1QTNF9*** | 412283 | 77 | 6942 | 5 | 3.95 (1.6, 9.79) | 0.00297 | 3.59 (1.81, 7.14) | 0.000261 |
| ***TMEM160*** | 412354 | 6 | 6945 | 2 | 20.1 (4.01, 101) | 0.000264 | 13.7 (3.28, 57.1) | 0.000328 |
| ***CHERP*** | 412358 | 2 | 6946 | 1 | 29.2 (2.59, 329) | 0.00636 | 36.5 (5.03, 264) | 0.000374 |
| ***PKP1*** | 412333 | 27 | 6943 | 4 | 8.88 (3.09, 25.5) | 4.91E-05 | 5.6 (2.15, 14.6) | 0.000428 |
| ***EML3*** | 412269 | 91 | 6939 | 8 | 5.38 (2.61, 11.1) | 5.33E-06 | 3.21 (1.68, 6.17) | 0.000447 |
| ***FOXP1*** | 412357 | 3 | 6945 | 2 | 31.7 (5.28, 190) | 0.000156 | 17.6 (3.54, 87.7) | 0.000456 |
| ***GBP2*** | 412281 | 79 | 6945 | 2 | 1.57 (0.384, 6.38) | 0.532 | 3.48 (1.73, 7.04) | 0.000494 |
| ***ASXL2*** | 412355 | 5 | 6946 | 1 | 12.2 (1.41, 106) | 0.0232 | 16.6 (3.4, 80.8) | 0.000512 |
| ***SNTG1*** | 412336 | 24 | 6943 | 4 | 9.97 (3.44, 28.9) | 2.22E-05 | 5.67 (2.08, 15.4) | 0.000683 |
| ***SLFN5*** | 411806 | 554 | 6944 | 3 | 0.33 (0.106, 1.03) | 0.0558 | 0.392 (0.228, 0.674) | 0.000701 |
| ***MOK*** | 383028 | 29332 | 6499 | 448 | 0.906 (0.823, 0.998) | 0.0458 | 0.904 (0.852, 0.958) | 0.000734 |
| ***DPP9*** | 412325 | 35 | 6943 | 4 | 6.73 (2.38, 19) | 0.000316 | 4.67 (1.91, 11.4) | 0.000738 |
| ***ELMOD2*** | 412076 | 284 | 6945 | 2 | 0.42 (0.105, 1.69) | 0.222 | 0.215 (0.0878, 0.525) | 0.000744 |
| ***UPF2*** | 412355 | 5 | 6946 | 1 | 11.1 (1.28, 95.9) | 0.0291 | 15.2 (3.12, 74) | 0.000748 |
| ***RARS2*** | 412096 | 264 | 6938 | 9 | 1.95 (1, 3.79) | 0.0496 | 2.14 (1.37, 3.32) | 0.000750 |
| ***RHO*** | 412290 | 70 | 6941 | 6 | 4.89 (2.12, 11.3) | 0.000196 | 3.4 (1.66, 6.93) | 0.000779 |
| ***TTR*** | 412352 | 8 | 6946 | 1 | 7.51 (0.933, 60.5) | 0.0581 | 11.2 (2.71, 46.7) | 0.000853 |
| ***JAGN1*** | 412036 | 324 | 6933 | 14 | 2.65 (1.55, 4.54) | 0.000363 | 2.02 (1.34, 3.06) | 0.000866 |
| ***NDOR1*** | 412189 | 171 | 6939 | 8 | 2.79 (1.37, 5.68) | 0.0046 | 2.45 (1.45, 4.16) | 0.000872 |
| ***CDCA3*** | 412309 | 51 | 6944 | 3 | 3.79 (1.18, 12.2) | 0.0253 | 4.07 (1.78, 9.33) | 0.000888 |
| ***TSC22D4*** | 412342 | 18 | 6946 | 1 | 3.55 (0.472, 26.7) | 0.219 | 7.15 (2.24, 22.8) | 0.000894 |
| ***FBXO42*** | 412357 | 3 | 6946 | 1 | 19.6 (2.01, 192) | 0.0105 | 21.6 (3.48, 133) | 0.000957 |
| ***FGF7*** | 412352 | 8 | 6946 | 1 | 7.35 (0.914, 59.1) | 0.0607 | 11 (2.65, 45.7) | 0.000959 |
| ***IK*** | 412220 | 140 | 6941 | 6 | 2.41 (1.06, 5.47) | 0.035 | 2.58 (1.47, 4.54) | 0.000960 |

### Supplementary Figures

#### Monte Carlo Simulations

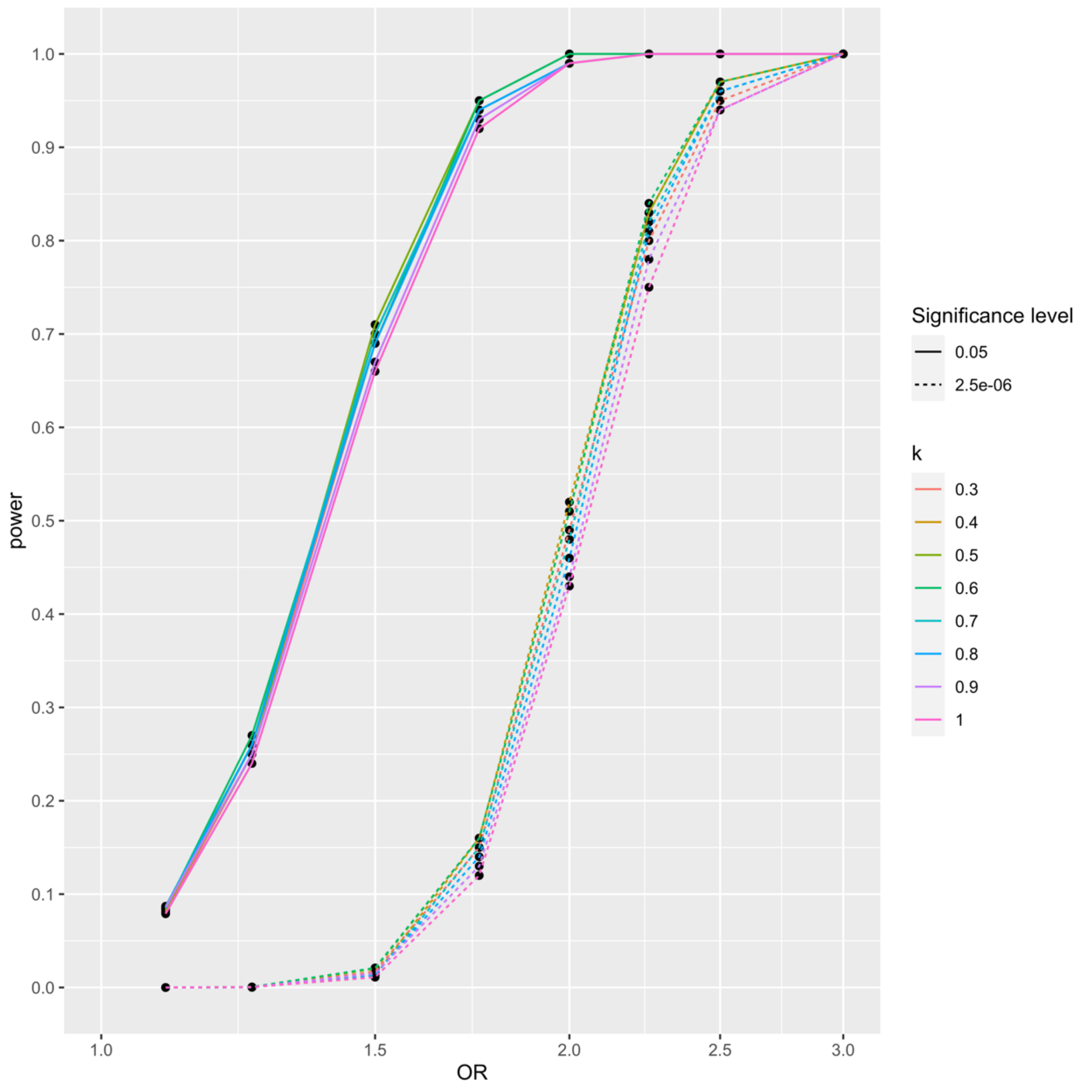

**Figure 1 | Comparing the power to detect different ORs at $\alpha = 0.05$ and at $\alpha = 2.5 \times 10^{-6}$ for varying k in model 3.** Power is calculated by Monte Carlo simulations for 5,000 datasets of size 450,000 with proportions of sex, breast cancer case/control, and family history the same as for each cancer in the UK Biobank. For each simulated dataset model 2 and model 3 were compared to the null model by LRT.

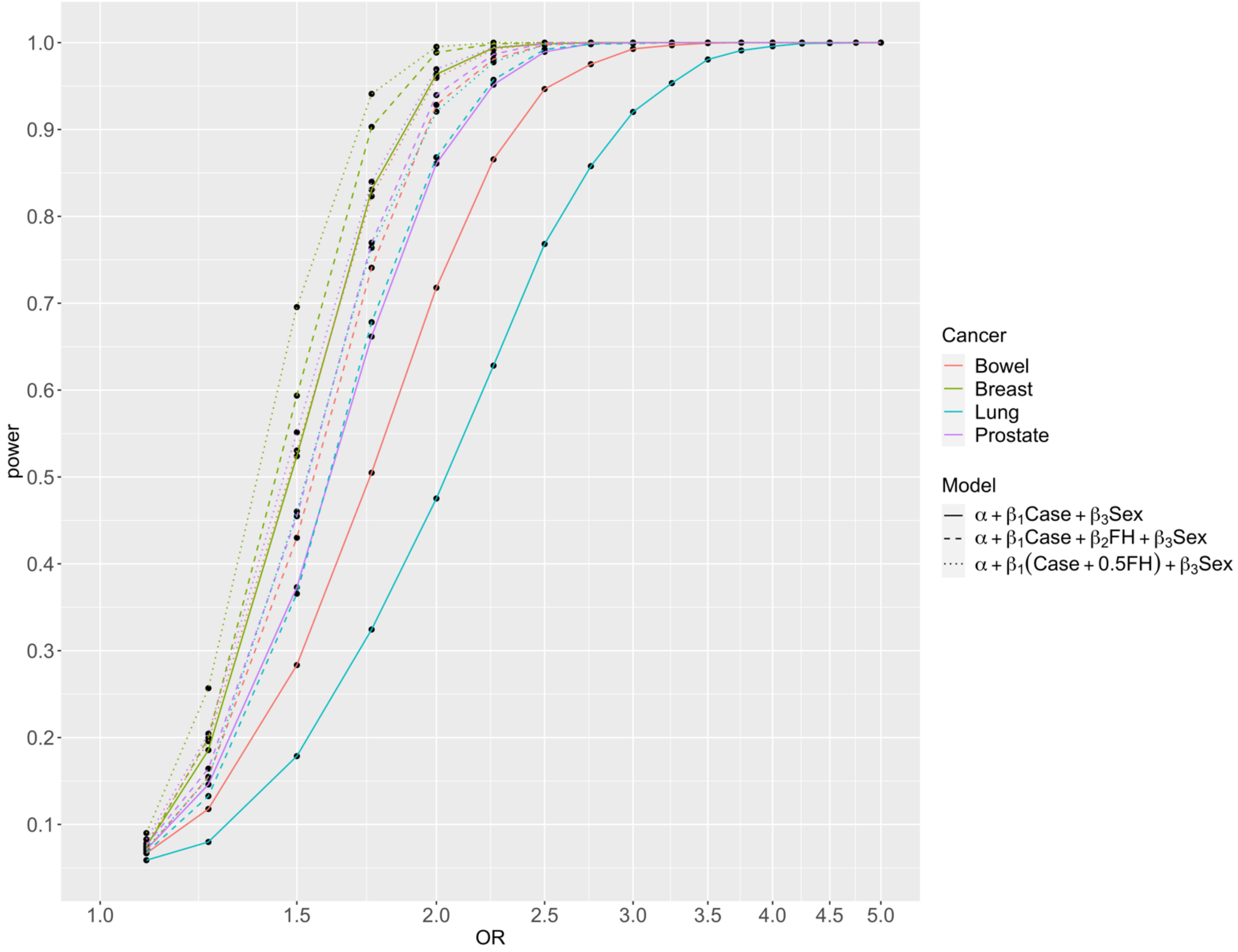

**Figure 2 | Comparing the power to detect different ORs at $\alpha = 0.05$ across cancers.** Power is calculated by Monte Carlo simulations for 5,000 datasets of size 450,000 with proportions of sex, case/control, and family history the same as for each cancer in the UK Biobank. For each simulated dataset model 2 and model 3 were compared to the null model by LRT.

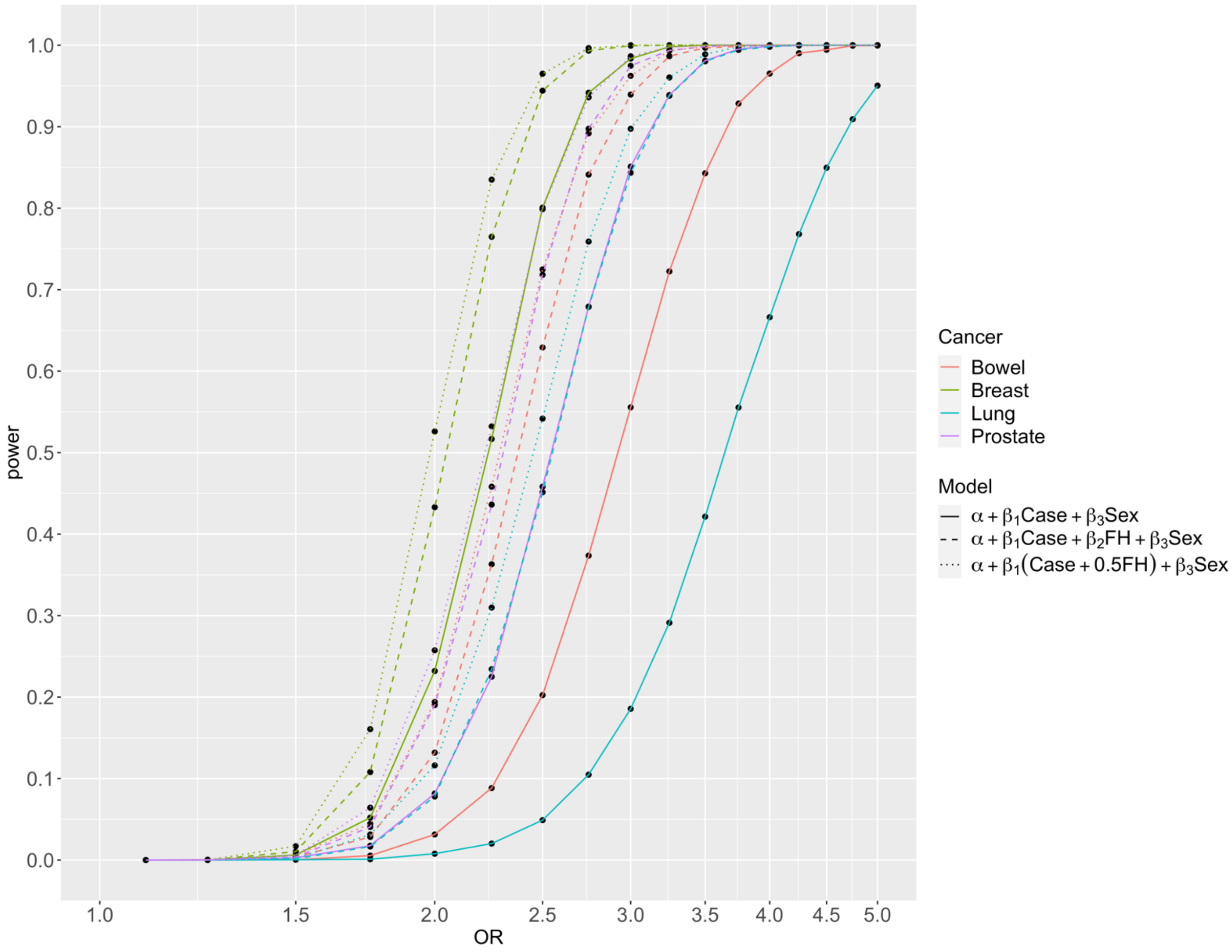

**Figure 3 | Comparing the power to detect different ORs at $\alpha = 2.5 \times 10^{-6}$ across cancers.** Power is calculated by Monte Carlo simulations for 5,000 datasets of size 450,000 with proportions of sex, case/control, and family history the same as for each cancer in the UK Biobank. For each simulated dataset model 2 and model 3 were compared to the null model by LRT.

#### Prostate Cancer

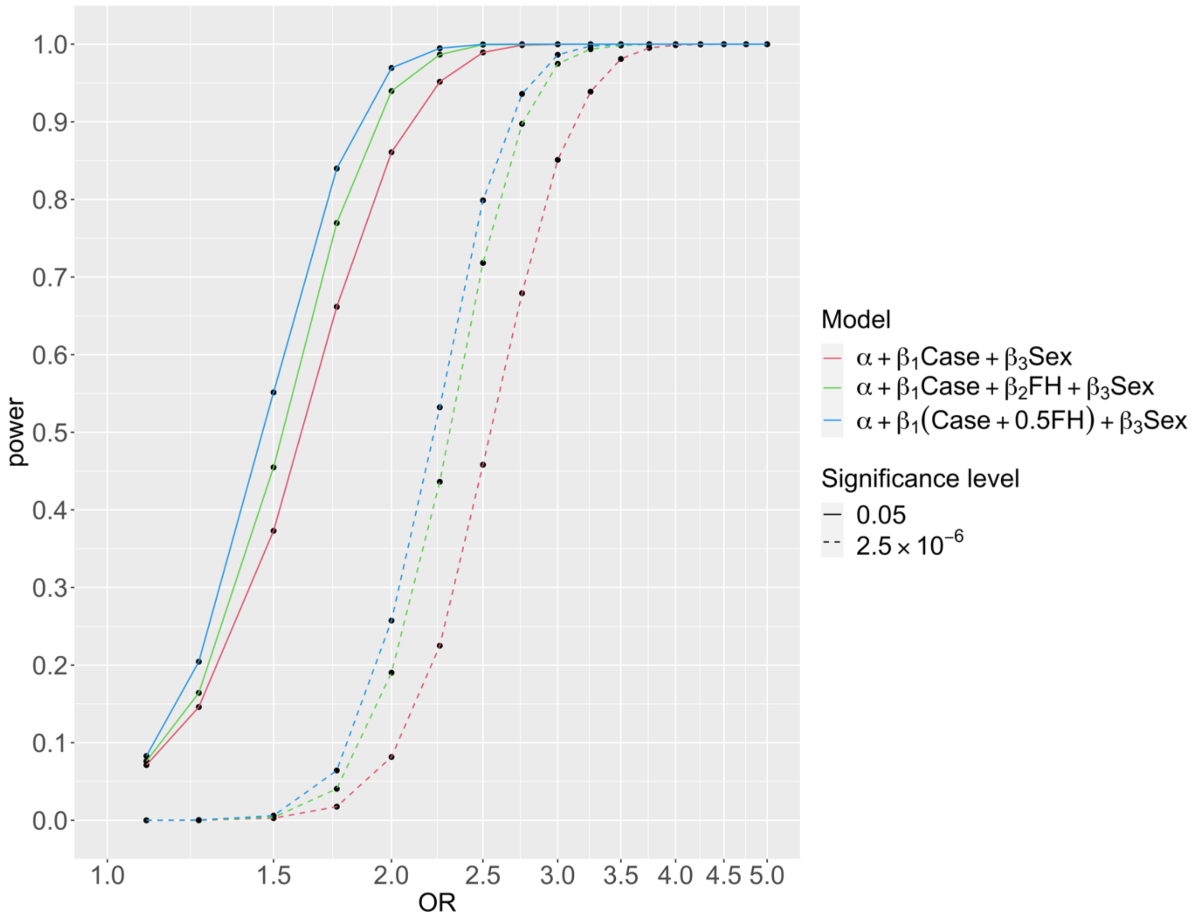

**Figure 4 | The power to detect different ORs for prostate cancer risk.** Power is calculated by Monte Carlo simulations for 5,000 datasets of size 450,000 with proportions of sex, case/control, and family history the same as for prostate cancer in the UK Biobank. For each simulated dataset model 2 and model 3 were compared to the null model by LRT.

Lung Cancer

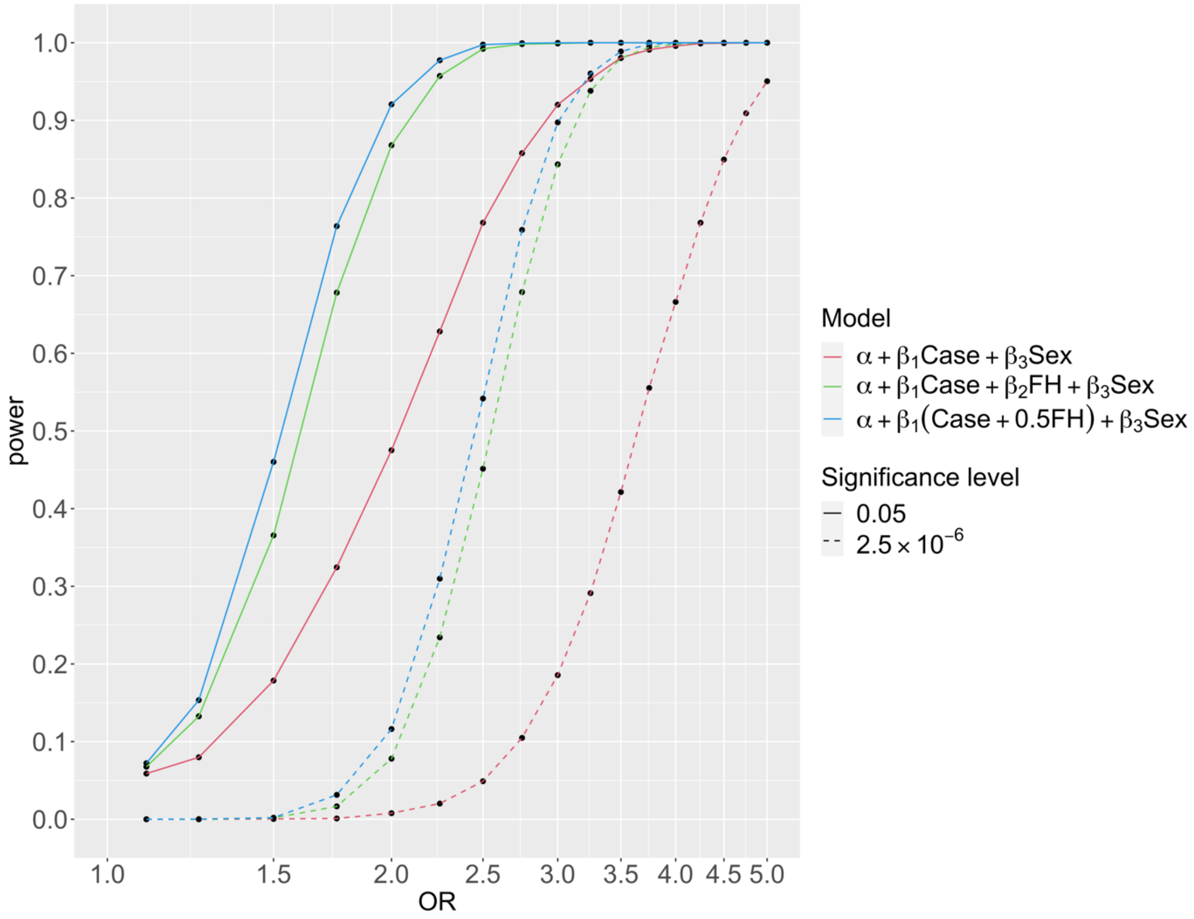

**Figure 5 | The power to detect different ORs for lung cancer risk.** Power is calculated by Monte Carlo simulations for 5,000 datasets of size 450,000 with proportions of sex, case/control, and family history the same as for lung cancer in the UK Biobank. For each simulated dataset model 2 and model 3 were compared to the null model by LRT.

Bowel Cancer

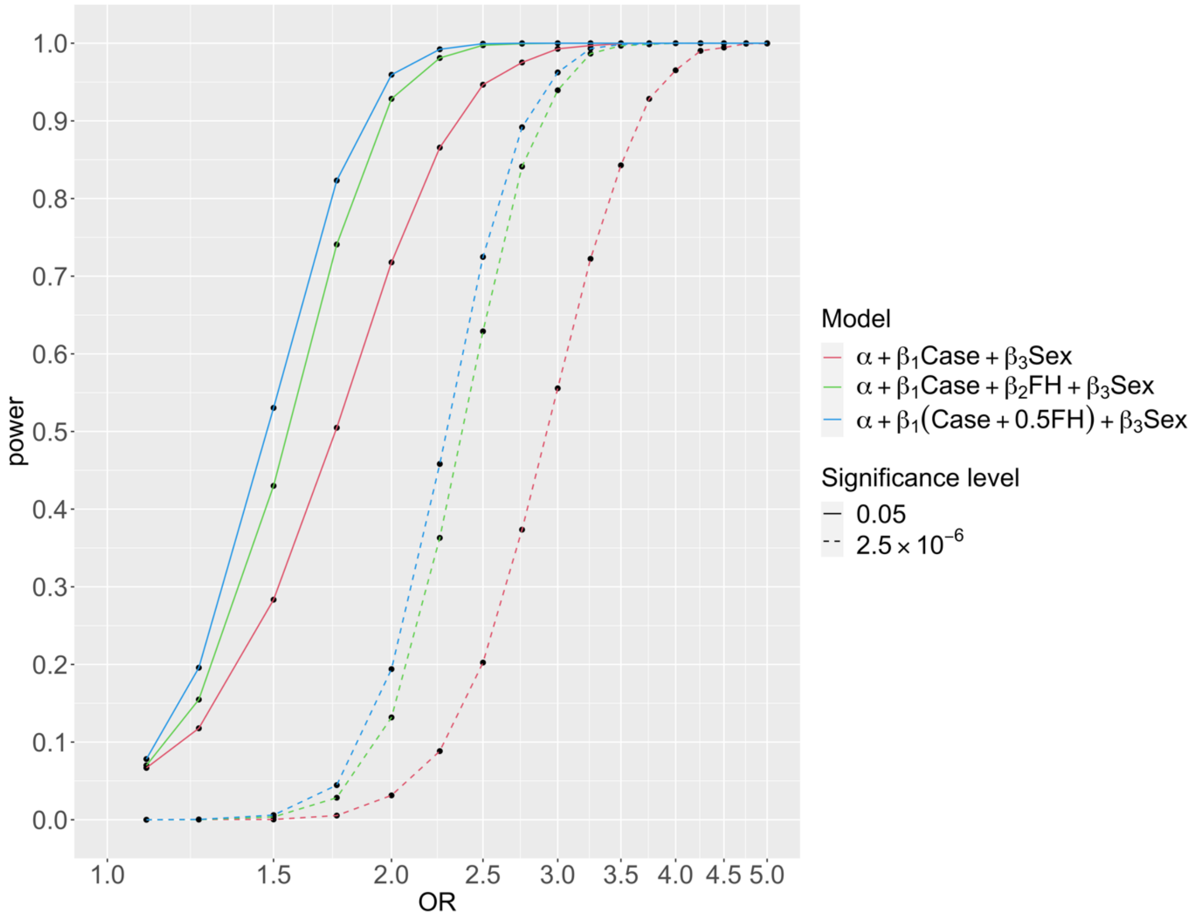

**Figure 6 | The power to detect different ORs for bowel cancer risk.** Power is calculated by Monte Carlo simulations for 5,000 datasets of size 450,000 with proportions of sex, case/control, and family history the same as for bowel cancer in the UK Biobank. For each simulated dataset model 2 and model 3 were compared to the null model by LRT.

Burden Test Results

Prostate Cancer

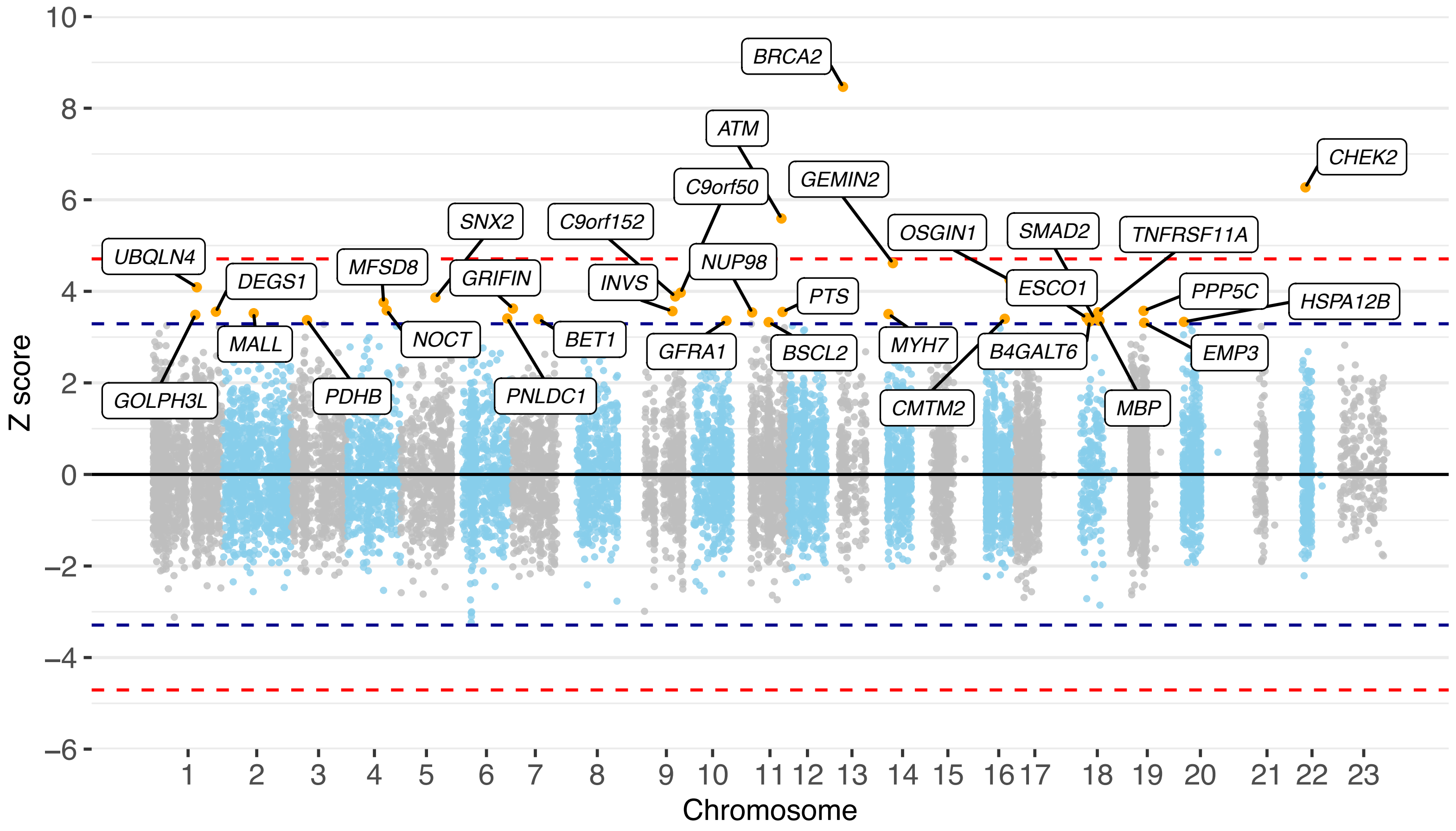

**Figure 7 |Manhattan plot of z scores from assessing the association between protein-truncating variant carriers within genes and prostate cancer risk, using model 3 with k=0.5.** The x axis is the chromosomal position, and the y axis is the z score from testing H0: β = ln(OR) = 0 (two-tailed) by LRT to the null model. The blue lines correspond to z = ±3.29, P = 0.001, the red lines correspond to z = ±4.71, P = 2.5 × 10−6. All labelled genes are those with P < 0.001. All P values are unadjusted for multiple testing.

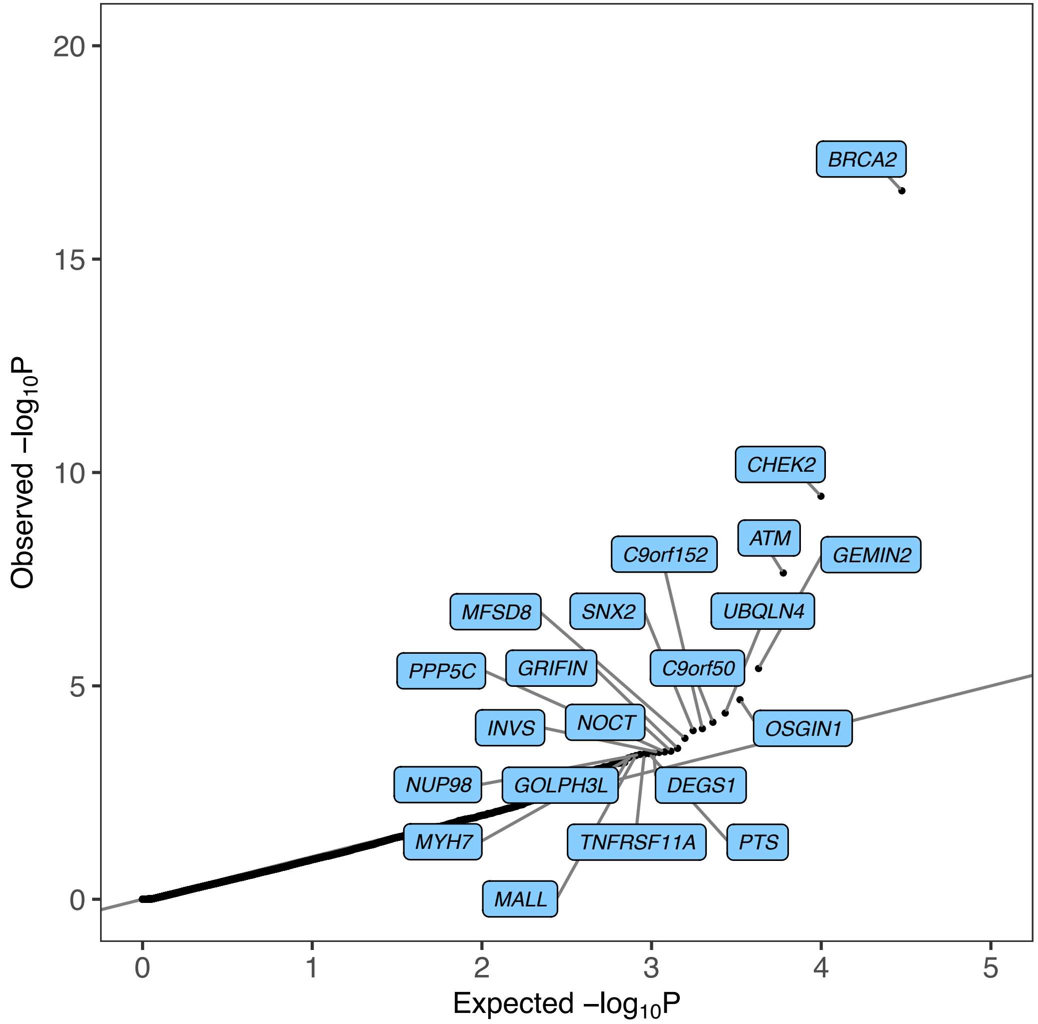

**Figure 8 | Quantile–quantile plot of P values from assessing the association between protein-truncating variant carriers and prostate cancer risk.** P-values are from testing H0: β = ln(OR) = 0 by LRT to the null model (two-tailed). The x axis is the expected log10 P values from the null hypothesis, the y axis is the observed log10 P values. All highlighted genes have P < 0.0005 and are associated with an increased risk of prostate cancer. All P-values are unadjusted for multiple testing.

Lung Cancer

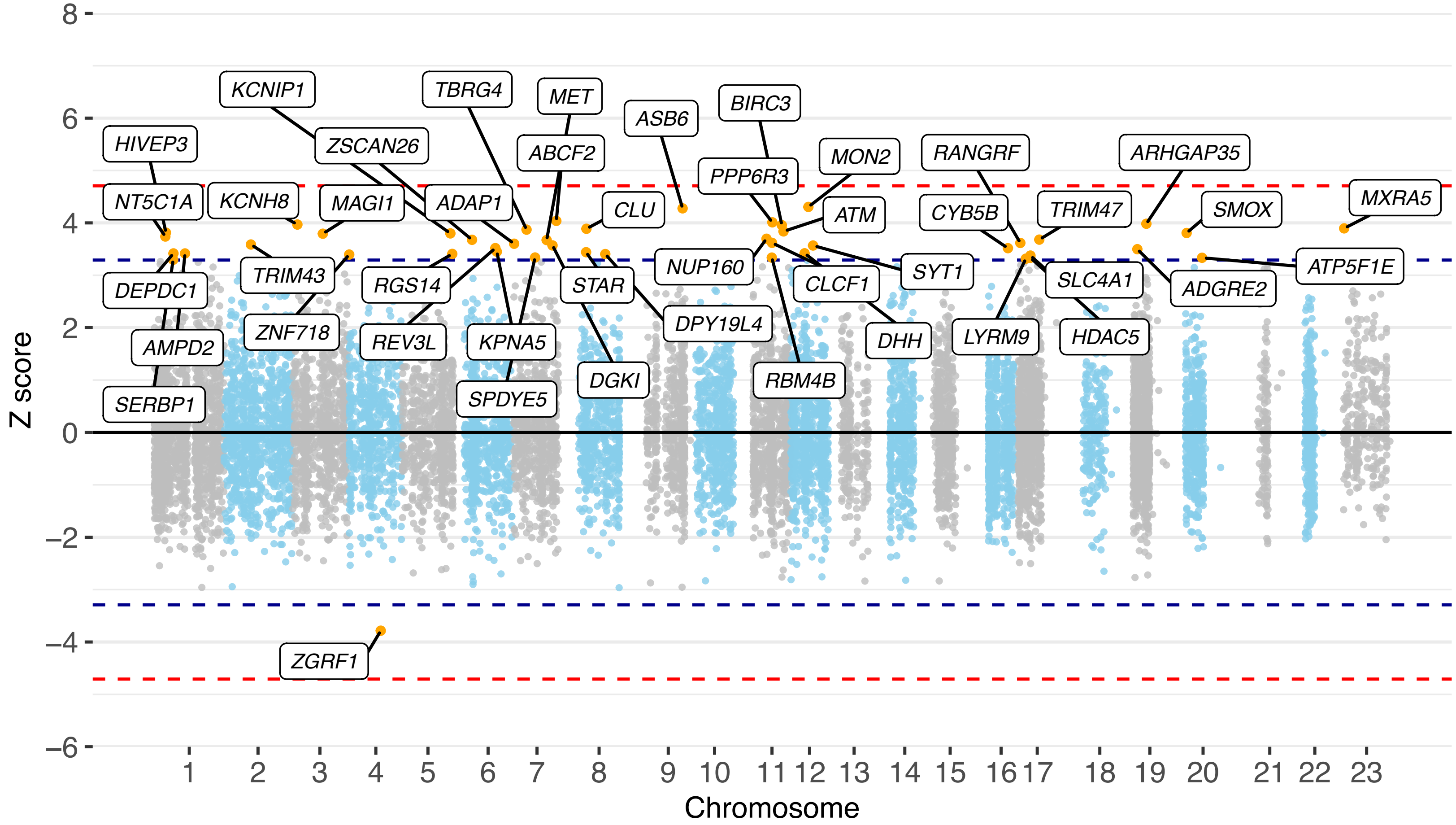

**Figure 9 | Manhattan plot of z scores from assessing the association between protein-truncating variant carriers within genes and lung cancer risk, using model 3 with k=0.5.** The x axis is the chromosomal position, and the y axis is the z score from testing H0: β = ln(OR) = 0 (two-tailed) by LRT to the null model. The blue lines correspond to z = ±3.29, P = 0.001, the red lines correspond to z = ±4.71, P = 2.5 × 10−6. All labelled genes are those with P < 0.001. All P values are unadjusted for multiple testing.

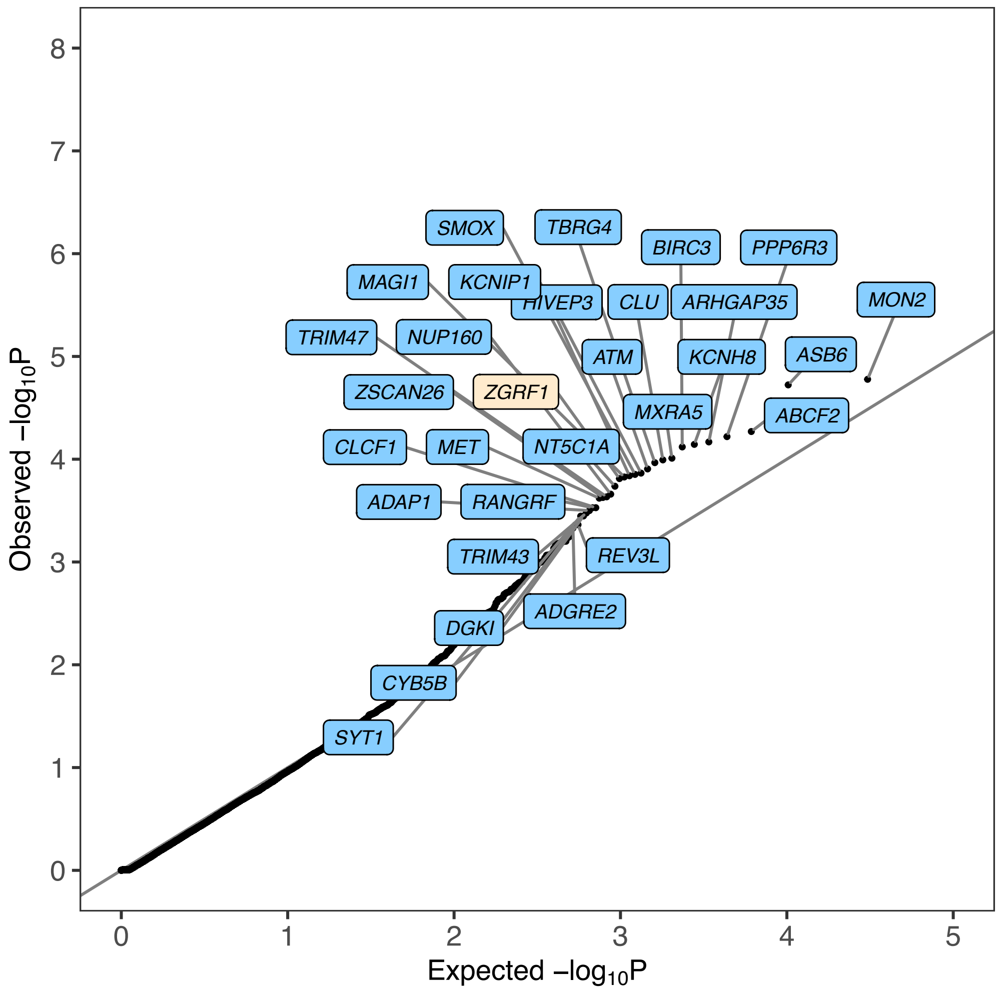

**Figure 10 | Quantile–quantile plot of P values from assessing the association between protein-truncating variant carriers and lung cancer risk.** P-values are from testing H0: $\beta = \ln(OR) = 0$ by LRT to the null model (two-tailed). The x axis is the expected log10 P values from the null hypothesis, the y axis is the observed log10 P value. Highlighted genes have $P < 0.0005$. Highlighted genes in blue are associated with an increased risk of lung cancer, and highlighted genes in cream are associated with decreased risk of lung cancer. All P-values are unadjusted for multiple testing.

Bowel Cancer

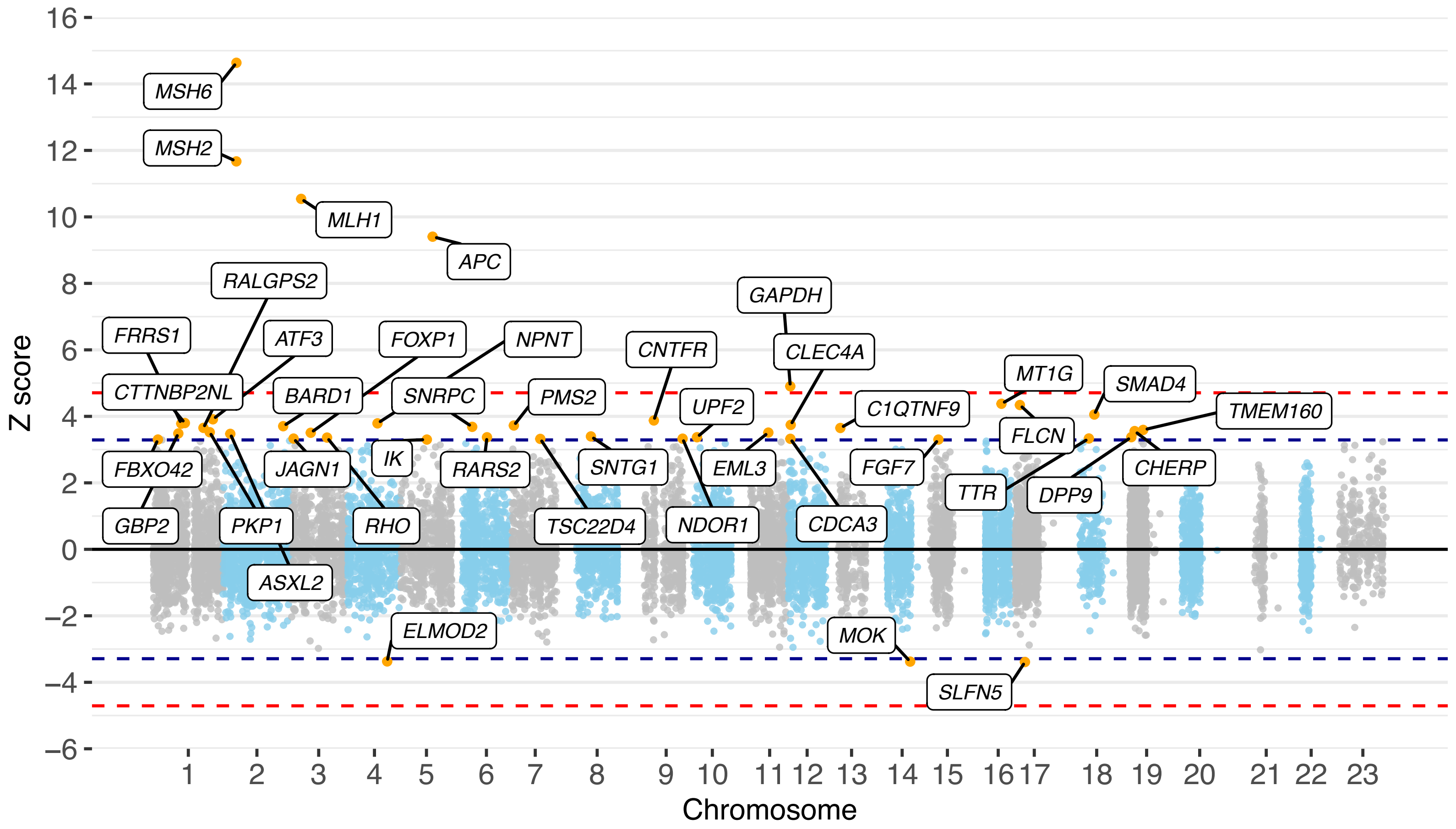

**Figure 11 | Manhattan plot of z scores from assessing the association between protein-truncating variant carriers within genes and bowel cancer risk, using model 3 with k=0.5.** The x axis is the chromosomal position, and the y axis is the z score from testing H0: $\beta = \ln(OR) = 0$ (two-tailed) by LRT to the null model. The blue lines correspond to $z = \pm3.29$, $P = 0.001$, the red lines correspond to $z = \pm4.71$, $P = 2.5 \times 10^{-6}$. All labelled genes are those with $P < 0.001$. All P values are unadjusted for multiple testing.

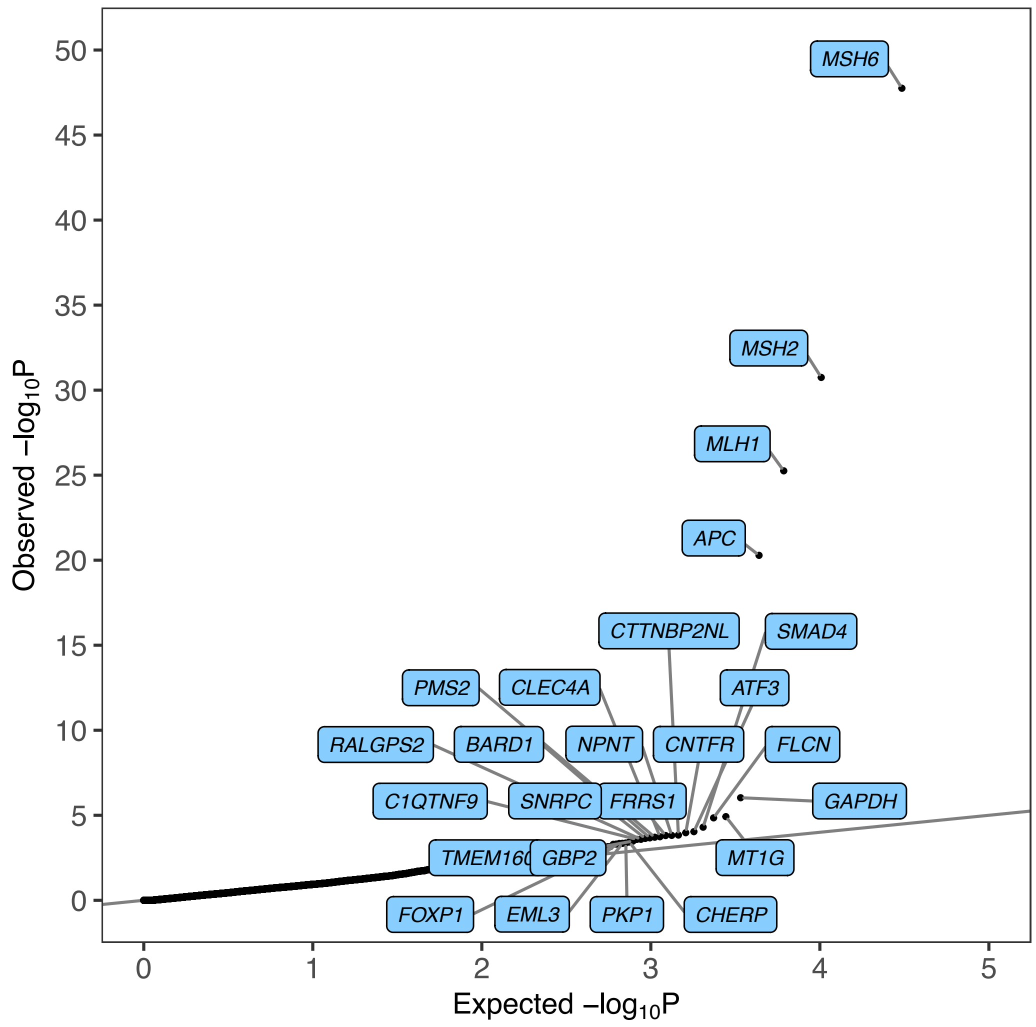

**Figure 12 | Quantile–quantile plot of P values from assessing the association between protein-truncating variant carriers and bowel cancer risk.** P-values are from testing H0: $\beta = \ln(OR) = 0$ by LRT to the null model (two-tailed). The x axis is the expected log10 P values from the null hypothesis, the y axis is the observed log10 P value. Highlighted genes have P < 0.0005. Highlighted genes in blue are associated with an increased risk of bowel cancer, and highlighted genes in cream are associated with decreased risk of bowel cancer. All P-values are unadjusted for multiple testing.
